# Modeling third waves of Covid-19 spread with piecewise differential and integral operators: Turkey, Spain and Czechia

**DOI:** 10.1101/2021.05.20.21257515

**Authors:** Abdon Atangana, Seda İğret Araz

## Abstract

Several collected data representing the spread of some infectious disease have demonstrated that the spread does not really exhibit homogeneous spread. Clear examples can include the spread of Spanish flu and Covid-19. Collected data depicting numbers of daily new infections in the case of Covid-19 from countries like Turkey, Spain show three waves with different spread patterns. A clear indication of crossover behaviors. While modelers have suggested many mathematical models to depicting these behaviors, it becomes clear that their mathematical models cannot really capture the crossover behaviors, especially passage from deterministic resetting to stochastics. Very recently Atangana and Seda have suggested a concept of piecewise modeling consisting in defining a differential operator piece-wisely, the idea was first in chaos and outstanding patterns were captured. In this paper, we extend this concept to the field of epidemiology with the aim to depict waves with different patterns. Due to the novelty of this concept, a different approach to insure the existence and uniqueness of system solutions are presented. A piecewise numerical approach is presented to derive numerical solutions of such models. An illustrative example is presented and compared with collected data from 3 different countries including Turkey, Spain and Czechia. The obtained results let no doubt for us to conclude that this concept is a new window that will help mankind to better understand nature.

## 1 Introduction

Although mankind has been using mathematical model to attempt replicating the spread patterns of infection diseases within given settlements, however in several cases, one will notice that their mathematical models do not always capture the wave and the different crossovers exhibiting by these waves. Some examples including the Spanish flu that started in 1918 and ended after three waves in 1920. Another recent one is the spread of Covid-19. The disagreement between experimental data and the solutions of mathematical equation is perhaps due to the fact that complexities of collected data are not sometimes taken into account. Another reason is perhaps the non-updating of existing theory, for example, many years the reproductive number have been used in many research papers with no new modification, indeed it is perhaps not worth to consider a reproductive number of a model describing spread that present three waves where each wave present a different pattern. Another example is the use of rates that have been always suggested to be constant, white a constant rate clearly show the homogeneity of the spread. Whereas in the normal situation rate of infection could be a function of time to indeed include into the mathematical equation the spread complexities or non-homogeneity. Another quit useful function to evaluate the stability of equilibrium point is the Lyapunov function. Such function cannot however inform about the wave and their patterns. Recorded data of Covid-19 spread of some countries show multiple waves. For example, collected data from Turkey, Czechia and Spain show three waves at least, where each wave present a different processes. Neither the concept of reproductive number or the Lyapunov function or the existing way of modeling epidemiological problems could possibly predict these waves. Indeed, modeling real world problems with crossover effects has always been a great challenge to modelers. Even though significant efforts have been made by researchers from different background, nevertheless there are still many real world problem exhibiting crossover behaviors with complex patterns. An effort made to address this resulted to an introduction of a fractional differential operators with non-singular kernel exhibiting a crossover from stretched exponential to power-law in terms of mean-square displacement. This non-singular kernel also has crossover from normal to sub-diffusion, also the kernel can express crossover from random walk to power-law. However, crossover from deterministic setting to stochastic or from stochastic to deterministic cannot be captured by this kernel, thus a great limitation of such operators to be used in this scenarios.

Very recently, Atangana and Seda suggested the concepts of piecewise differential and integral operators. Additionally, the extended their innovative idea to modeling by suggesting a piecewise modeling. This idea is perhaps the future of modeling with this innovative idea, we suggest in this paper a new way to model epidemiological problems exhibiting crossover behaviors. In this paper, two illustrative examples will be presented. We will assume that real world spread exhibit three waves with different patterns, classical, nonlocal and randomness, a permutation can be done to have different behaviors based on the above processes.

## 2 Modeling spread of infectious diseases with waves

In this section, we present a new way to model spread of infectious diseases exhibiting waves with different patterns. Let assume without loss of generality that a mathematical model with n classes describes spread of an infectious diseases with three waves where each wave exhibits different patterns, assume with no loss of generality that classical mechanical processes, non-local processes, randomnessly and their permutation, which will be considered in different cases.

### 2.1 Case 1: Classical-power-law-randomness

In this case, we assume a scenario where the three waves of the spread exhibit the above processes. We assume that the first process starts from 0 to *T*_1_, the second from *T*_1_ to *T*_2_, and the last from *T*_2_ to *T*. A piecewise mathematical model associate to this can be given as

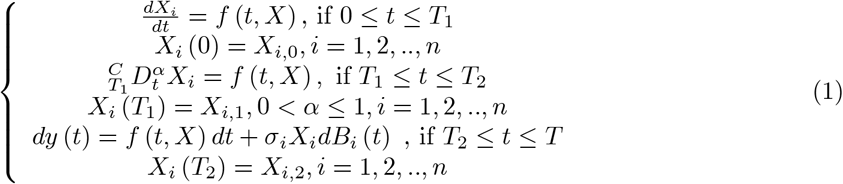

where *σ*_*i*_ are densities of randomness and *B*_*i*_ are the functions of noise.

### 2.2 Case 2: Classical-Mittag-Leffler law-randomness

In this case, we assume a scenario where the three waves present processes with classical behaviors, passage from stretched exponential to power-law and randomness, within the intervals described earlier in case (1). A piecewise model associate to the above scenarios can be given as

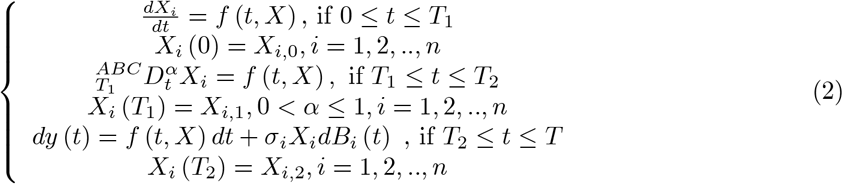

### 2.3 Case 3: Classical-fading memory-randomness

In this case, it is assumed that three waves are observed where the second wave displays fading memory process, which was known to be described best with the differential operator with exponential kernel. We also assume that three waves appear in the intervals described in case (1), then, the mathematical model associate to this scenario is given as:

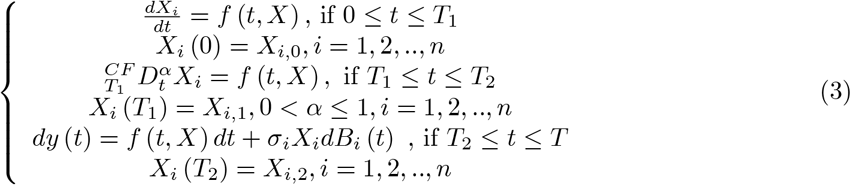

It is noted that in case (1), (2) and (3)

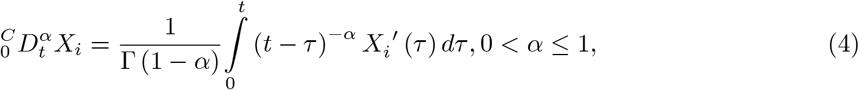

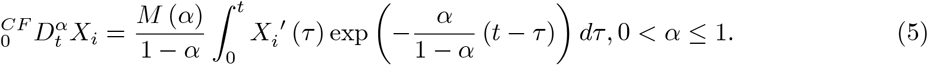

where *M* (*α*) is the normalization function such that *M* (0) = *M* (1) = 1. Also

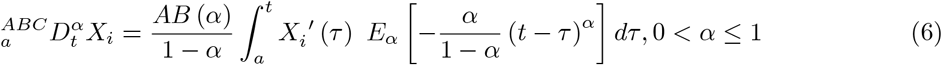

where 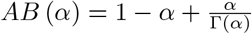 is the normalization function such that *AB* (0) = *AB* (1) = 1.

The analysis regarding the theory of existence and uniqueness will not be discussed here in general. However, this can be achieved within each interval In our paper, we will present existence and uniqueness for some examples. But here, we shall present numerical solution of such model.

## 3 Numerical solution of piecewise epidemiological model

Assuming that the described models satisfy theoretical aspects of existence and uniqueness. Thus in this section, we present numerical solutions. We shall use in all cases based on the Newton polynomial interpolation.

### 3.1 Numerical method for Case 1

We consider the first case

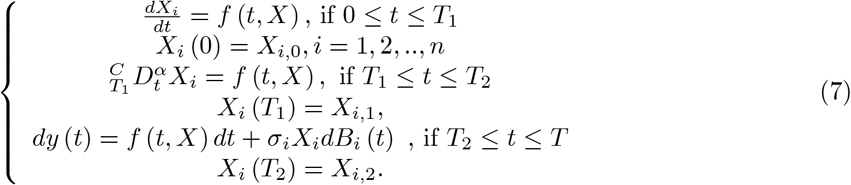

We divide [0, *T*] in three

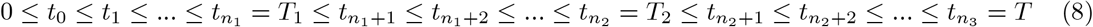

The numerical solution can then be provided as

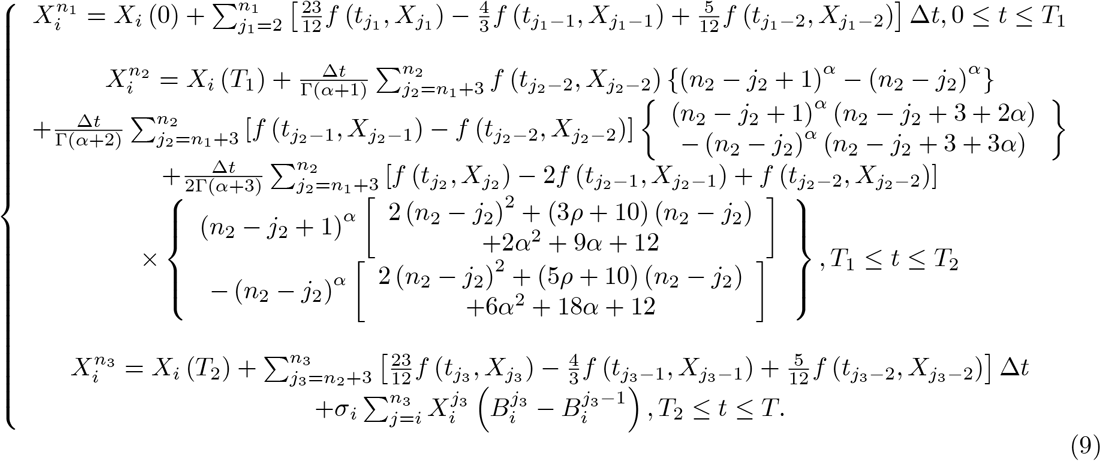

### 3.2 Numerical method for Case 2

We deal with the following problem with second case

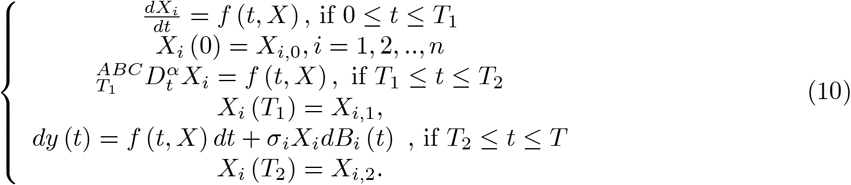

The numerical solution for such problem is given by

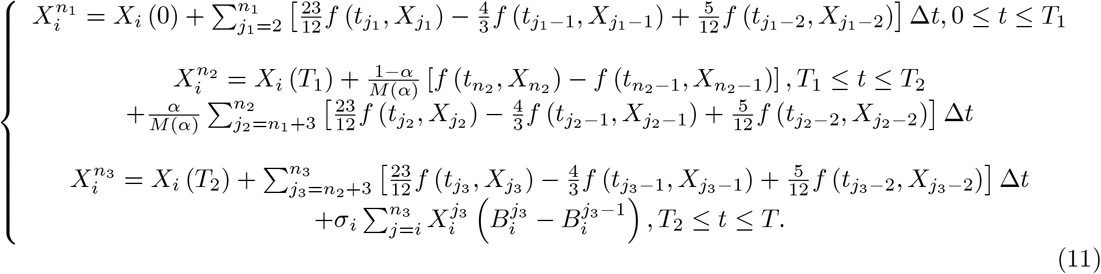

### 3.3 Numerical method for Case 3

We consider the following problem with third case

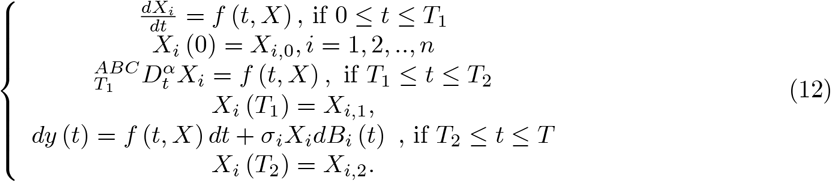

Using same routine, the numerical solution can be obtained as

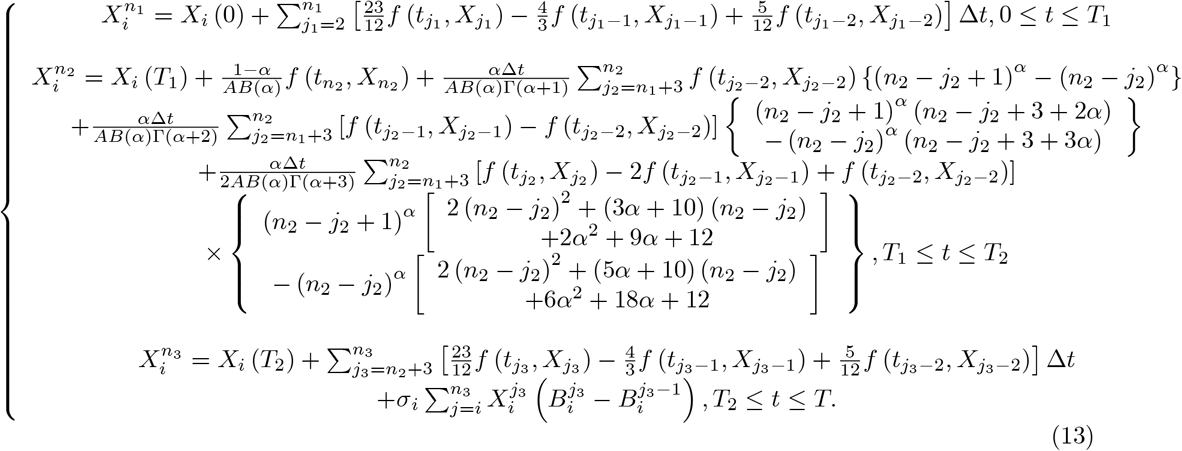

## 4 A mathematical model of Covid-19 spread with piecewise modeling

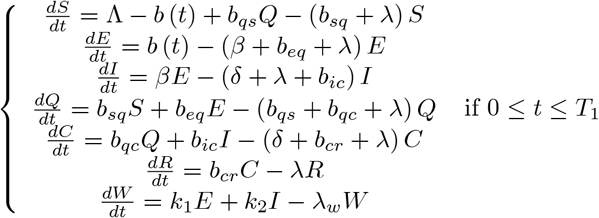

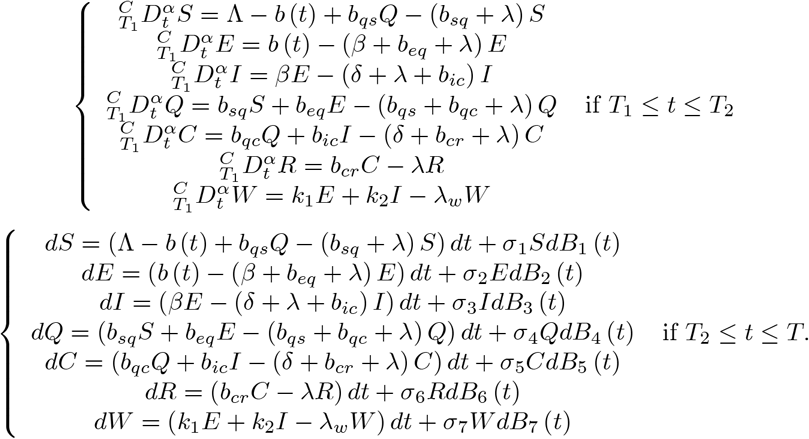

where

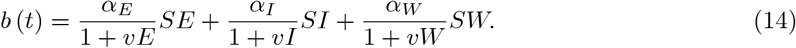

The function *S* (*t*) is the number of individuals susceptible to infection at a given time *t*. The function *E* (*t*) is the number of individuals infected but without symptoms at a given time. The function *I* (*t*) is the number of individuals infected but not yet isolated at a given time *t*. The function *Q* (*t*) is the number of individuals quarantined at a given time *t*. The function *C* (*t*) is the number of individuals confirmed to be infected with the virus, isolated and expecting recovery at a given time. The function *R* (*t*) is the number of individuals recovered at a given time *t*. The function *W* (*t*) is the concentration of the virus in the environmental reservoir. The initial conditions

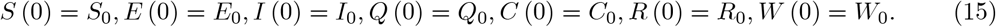

**Table 1.** Parameters of the suggested Covid-19 model

|  |  |
| --- | --- |
| $\Lambda$ | : Birth rate |
| $\lambda$ | : Natural mortality rate |
| $\delta$ | : The disease-induced fatality rate |
| $\beta^{-1}$ | : The incubation period between the infected and the onset of symptoms |
| $b_{cr}$ | : The rate of recovery from the infectious disease |
| $b_{ic}$ | : The rate at which highly infectious individuals are confirmed |
| $b_{sq}$ | : The rate at which susceptible are quarantined |
| $b_{eq}$ | : The rate at which exposed are quarantined |
| $b_{qc}$ | : The rate at which quarantine individuals are confirmed |
| $b_{qs}$ | : The rate at which quarantined move back to the susceptible class |
| $k_1$ | : The rate at which the exposed contribute the disease to the environmental reservoir |
| $k_2$ | : The rate at which the infected contribute the disease to the environmental reservoir |
| $\lambda_w$ | : Rate of removal of the virus from the environmental reservoir |
| $N$ | : Total population |
| $\alpha_E$ | : Transmission rate from the exposed to the susceptible |
| $\alpha_I$ | : Transmission rate from the highly infected to the susceptible |
| $\alpha_W$ | : Transmission rate from the environmental reservoir to the susceptible |
| $v$ | : Coefficient providing adjustment to the transmission rate |

## 5 Existence and uniqueness of Covid-19 model

In this section, we attempt to present a proof of existence and uniqueness of system solution of this model. However, we shall note that, different techniques will be applied in respect to each interval, indeed for the first and second case, the well-known procedure using the Banach fixed-point theorem or modified versions will be applied, however, the last part, since concerning stochastic a different technique is applied.

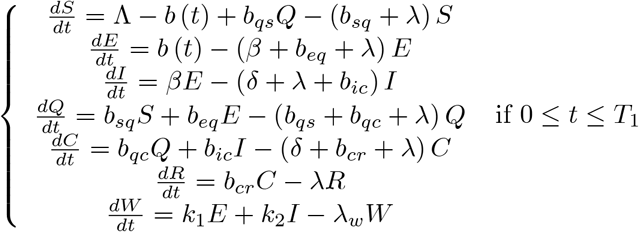

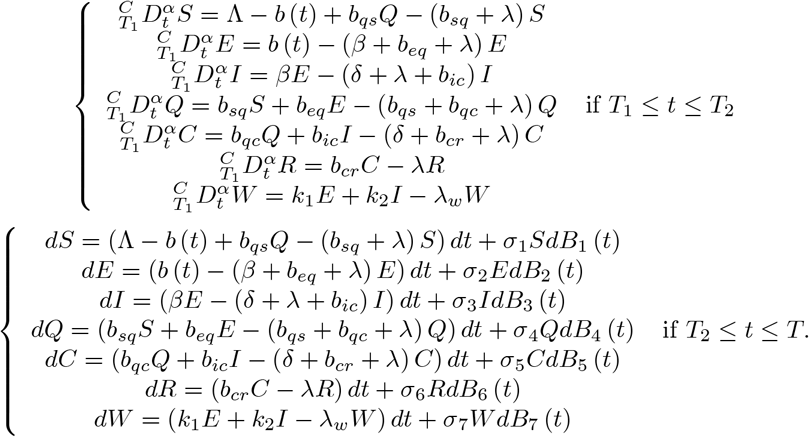

We now present the existence and uniqueness of the piecewise model. For simplicity, we write subject to *X* = (*S, E, I, Q, C, R, W*)

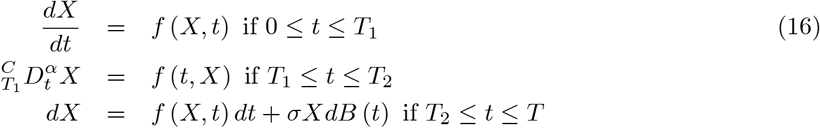

where

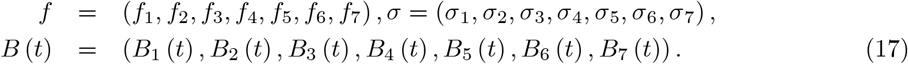

To show the existence and uniqueness, we shall prove that ∀ [0, *T*_1_] and [*T*_1_, *T*_2_], *f*_*i*_ (*X, t*) satisfy

1. Linear growth condition
2. The Lipschitz condition.

∀ *t* ∈ [*T*_2_, *T*], another methodology will be adopted.

For proof, we consider for ∀ *t* ∈ [*T*_2_, *T*].

Since all the used parameters of our model a positive constant, thus, they are continuous in Lipschitz sense for a given initial size of population 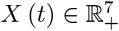, one can find a unique local solution. *X* (*t*) ∈ [*T*_2_, *τ*_*e*_), where *τ*_*e*_ denotes explosion time. To insure the solution is global, one has to prove that such system solution is global, we should prove that *τ*_*e*_ = ∞. To achieve our goal, we consider *k*_0_∈ ℝ _+_ is a positive constant such that *X* (*T*_2_) lies within 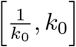 Additionally, we define a stopping time

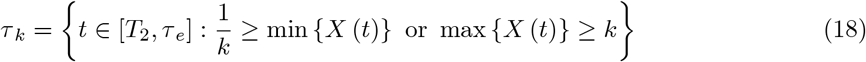

for each *k* ≥ *k*_0_.

*τ*_*k*_ is monotonically increasing as *k* → ∞. lim _*k* → ∞_ *τ*_*k*_= *τ*_*∞*_ with *τ*_*e*_ ≥ *τ*_∞_. ∀ *t* ≥ 0,if we show that *τ*_∞_= 0, then we can conclude that *τ*_*e*_ = → and 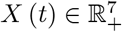 is solution. To achieve that we have to prove that *τ*_*e*_ = ∞. Nevertheless if the conclusion is contradictory, then there exists 0 < *T* and *ξ* ∈ (0, 1) such that

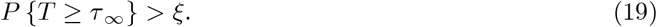

We additionally define a function 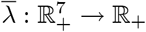 in *λ* ∈ *C*^2^ space such that

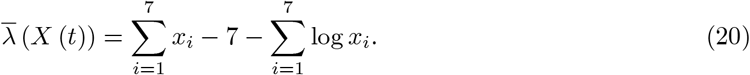

Owning the fact that ∀*z* > 0, *z* - 1 - log *z* ≥ 0, this leads to 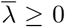. Additionally, it is assumed that *k*_0_ <*k* and 0 < *T*, therefore a direct application of Ito formula leads to

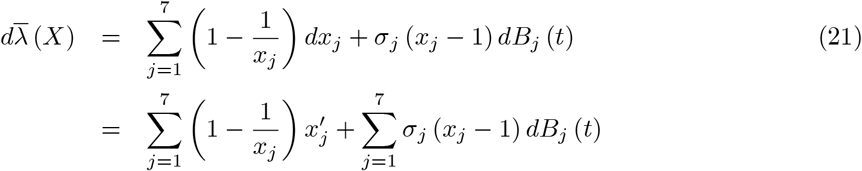

Here

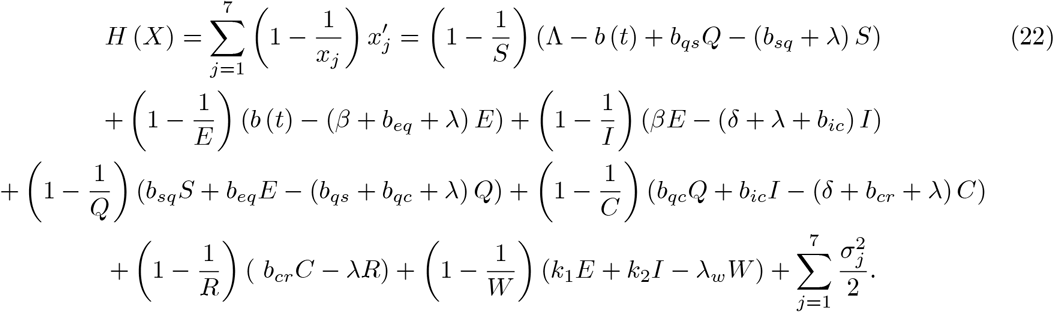

Then

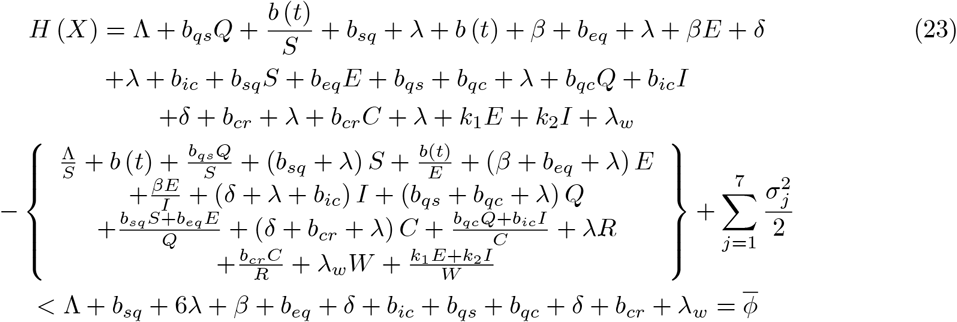

and

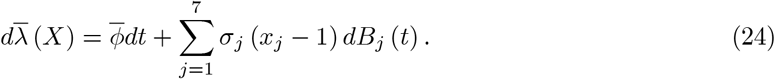

By direct integration from 0 to *τ*_*k*_ ∧*T*, we have

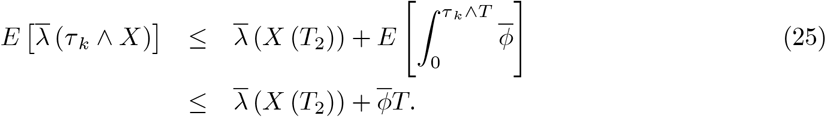

Setting Π _*k*_ = {*T* > *τ*_*k*_ } for *k*_1_≤ *k* and thus *p* (Π _*k*_) ≥ *ξ*. Noting that for ∀ Ω ∈Π _*k*_, there must exist at least one *X* (*τ*_*k*_, *w*) which is equal to 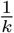 or *k*. Then *k* −log *k*−1 or 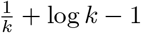 as result

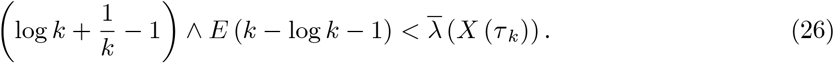

From above, we can write

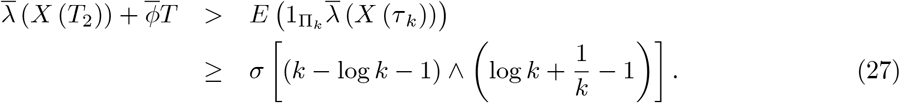

Here 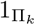 is the indicator function of Π. Thus lim_*k* → ∞_ leads

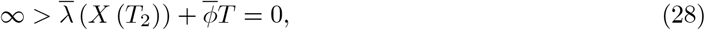

which is a contradiction. Therefore under the conditions presented earlier *τ*_∞_ = ∞ which completes the proof. Within [0, *T*_2_] we show that ∀ [0, *T*_2_], *i* = 1, 2,.., 7

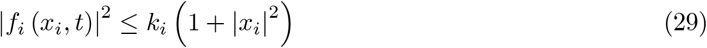

and

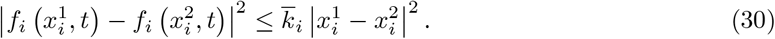

For ∀ ∈ [0, *T*], we have

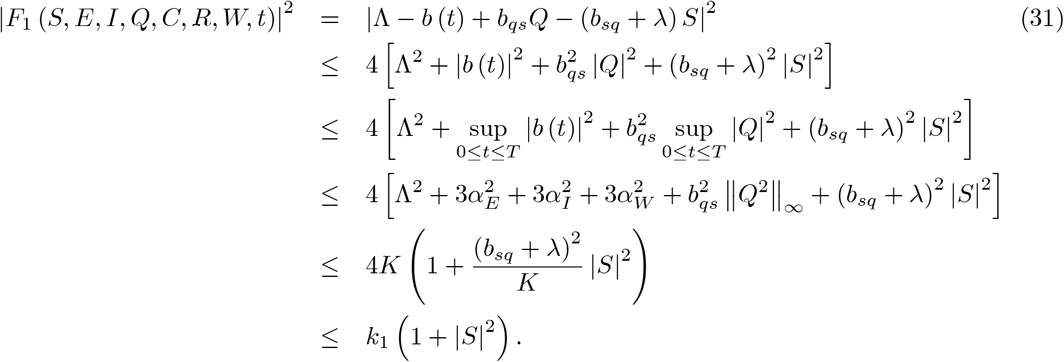

where 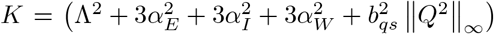 and under the condition 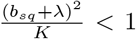. For ∀ *t* ∈ [0, *T*], we get

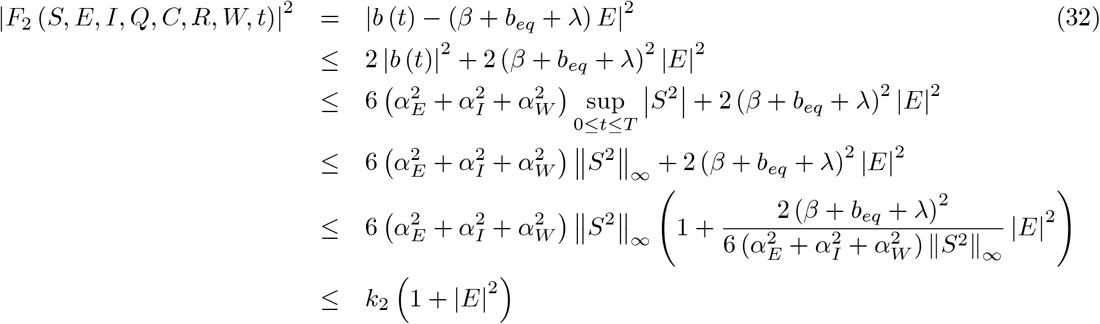

if 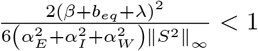.

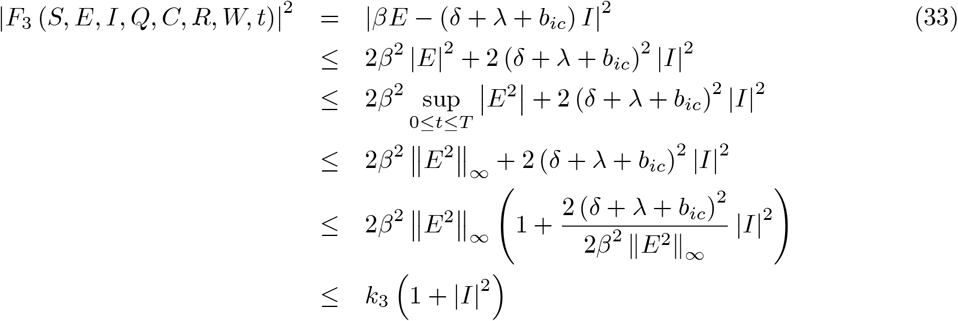

if 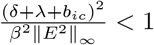.

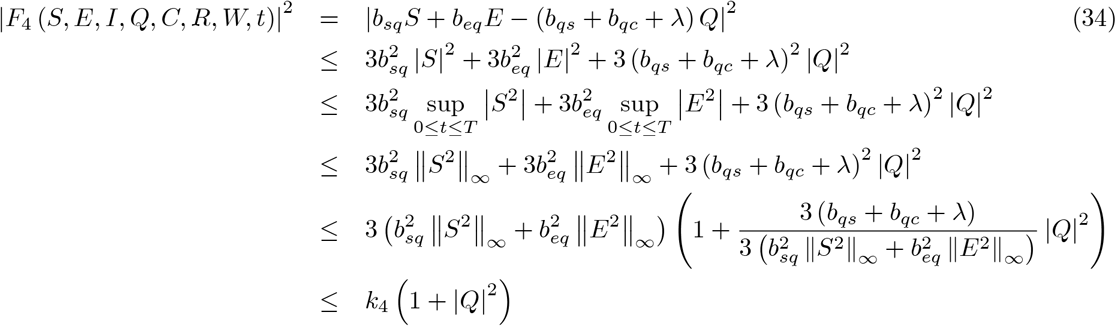

under the condition 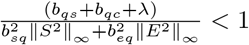.

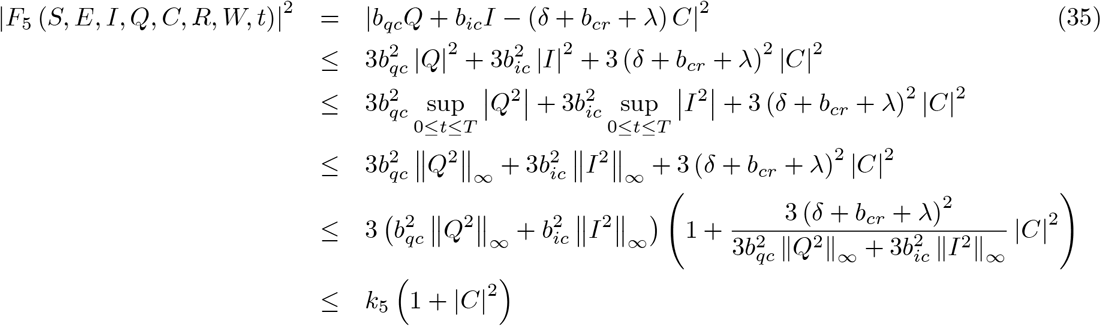

if 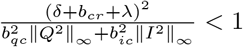.

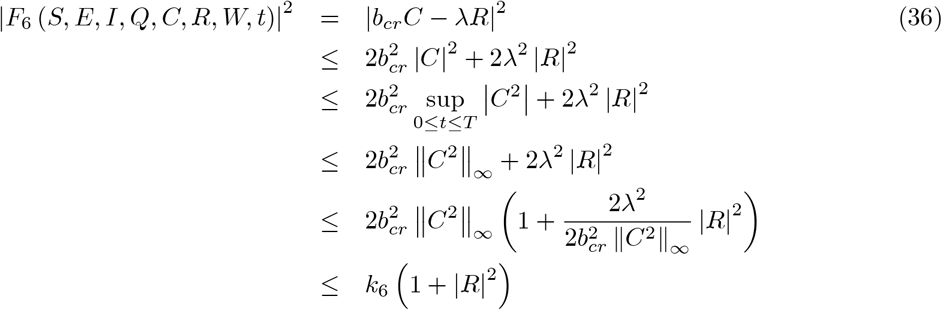

if 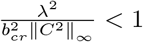. Finally, we have

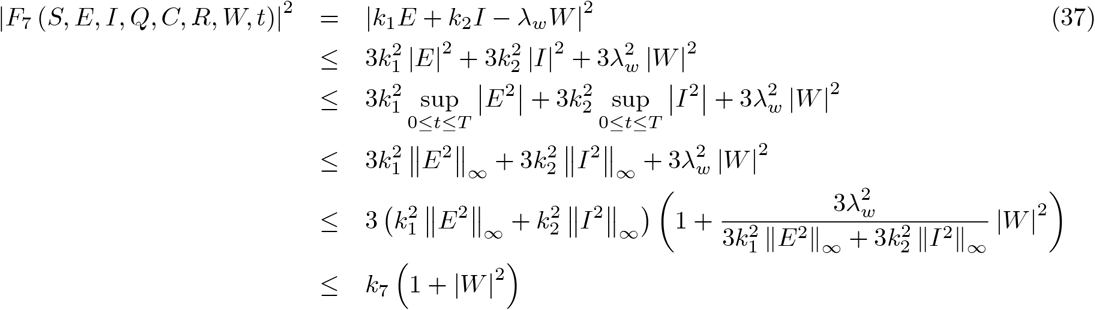

under the condition 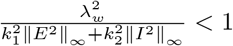. To verify second condition, we write

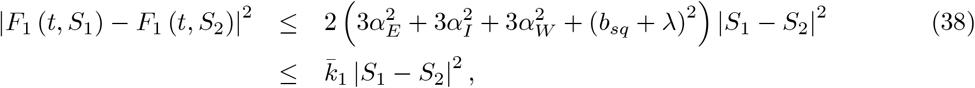

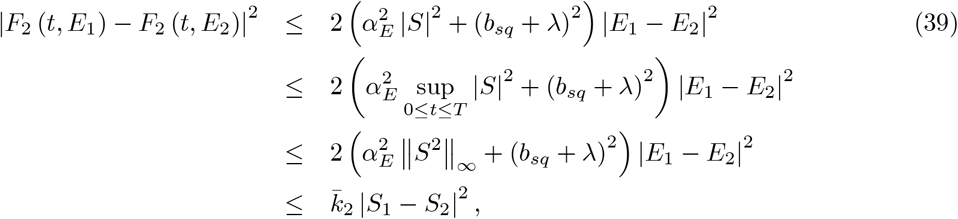

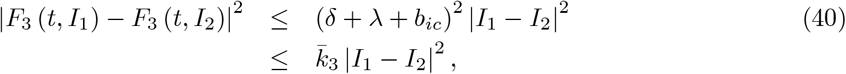

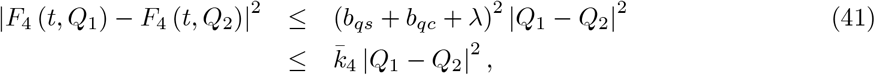

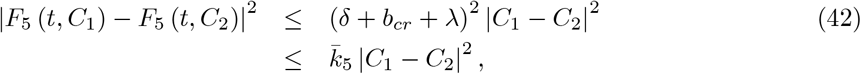

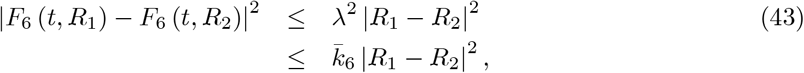

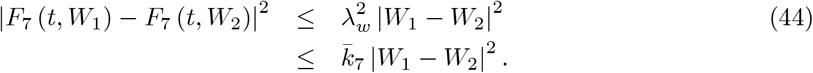

Therefore under the condition that

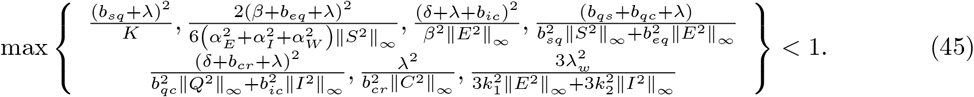

Then assumed that the solution are positive in [0, *T*_2_], then ∀ *t* ∈ [0, *T*] the piecewise has a unique positive solution.

## 6 Applications for Covid-19 model

A demonstrative example is better than precept, therefore, in this section, we present an application of the suggested theory. In particular, we will consider a mathematical model that was suggested to depicting the spread of Covid-19 within a given settlement. The model will be later modified to follow the steps presented in piece-wise modeling and for each model, a numerical method will be used to provide a numerical solution to the model. Numerical simulation using the suggested numerical are performed and depicted in Figure 1 to 7 for the first case, Figure 8 to 21 for second case and lastly Figure 22 to 28 for third case.

### 6.1 Numerical simulation of Covid-19 model for Case 1

In case one, we consider a country within which the spread displays three processes, including classical behaviors, power law behavior and finally stochastic behaviors. In this case, if we consider *T* as the last time of the spread, this is to say, the last day where a new infection occur, then, for the first period of time ranging from 0 to *T*_1_, the mathematical model will be constructed with classical differential operator, the second phrase, the model will be with the Caputo-Power law differential operator and finally stochastic approach will be used at the last phase. The mathematical model explaining this dynamic is then presented below.

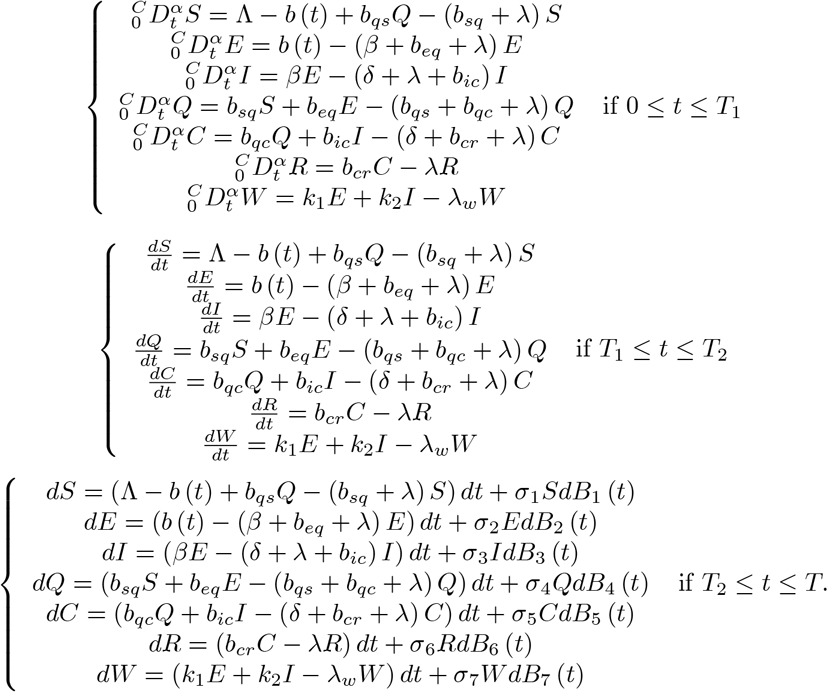

The initial conditions are considered as

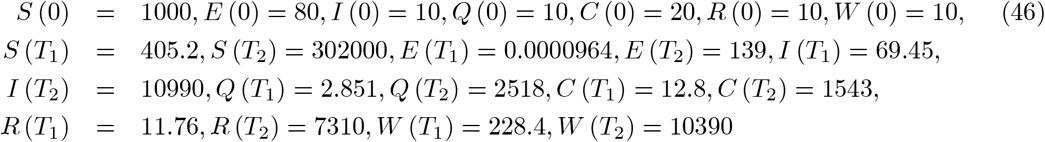

and the parameters are

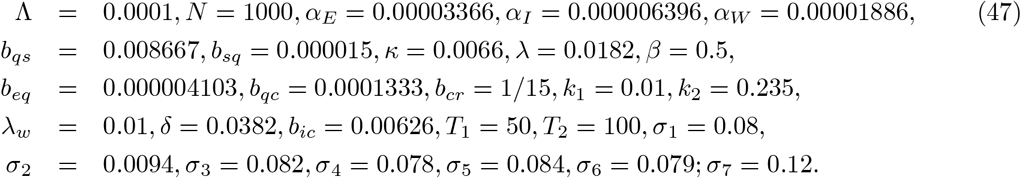

The numerical simulations for piecewise model are performed in Figure 1-7.

**Figure 1.**
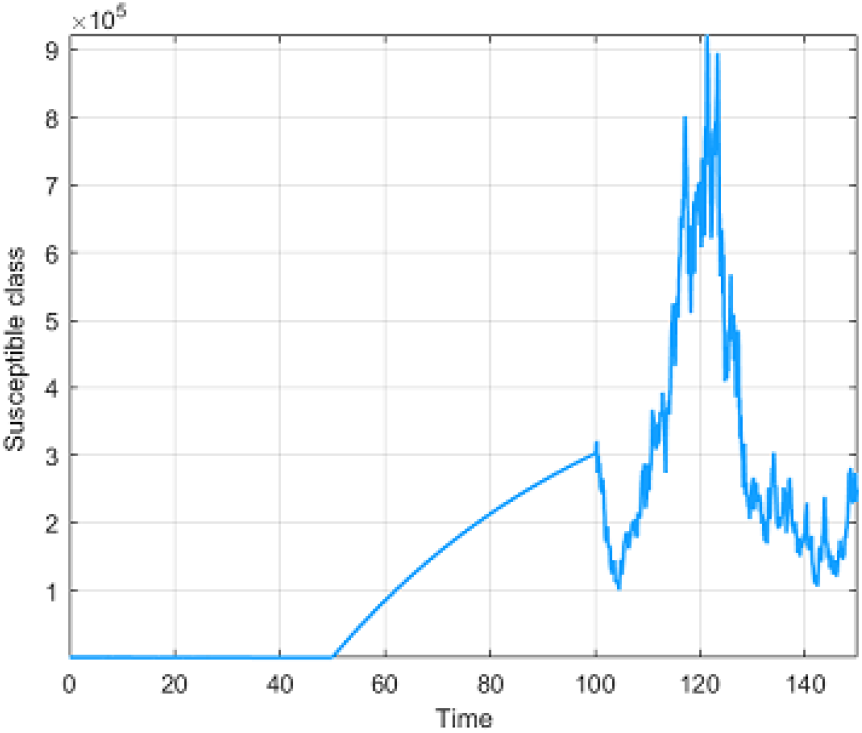
Numerical visualization for susceptible people for *α* = 1.

**Figure 2.**
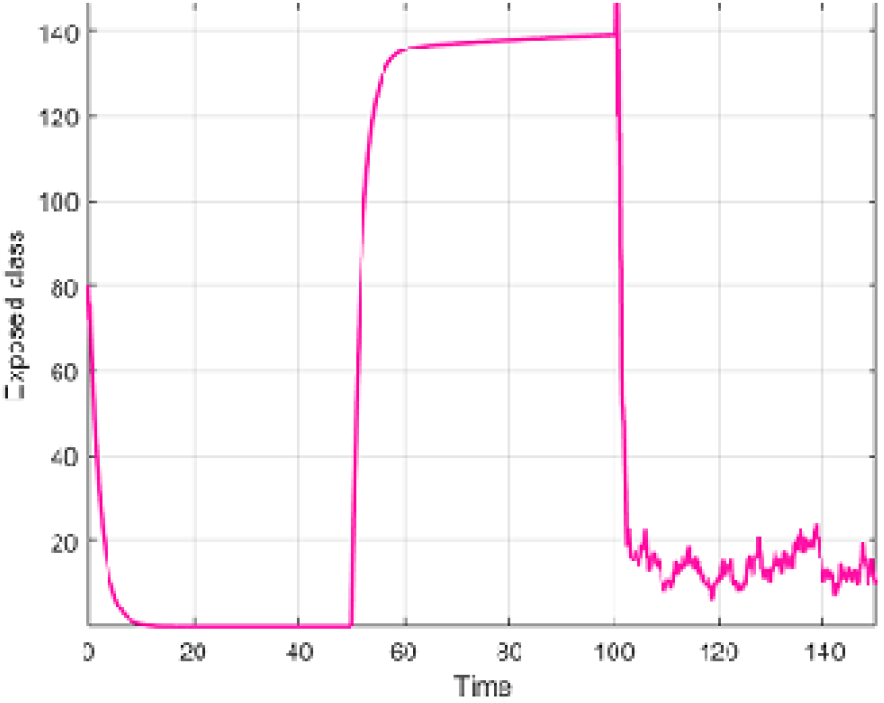
Numerical visualization for exposed people for *α* = 1.

**Figure 3.**
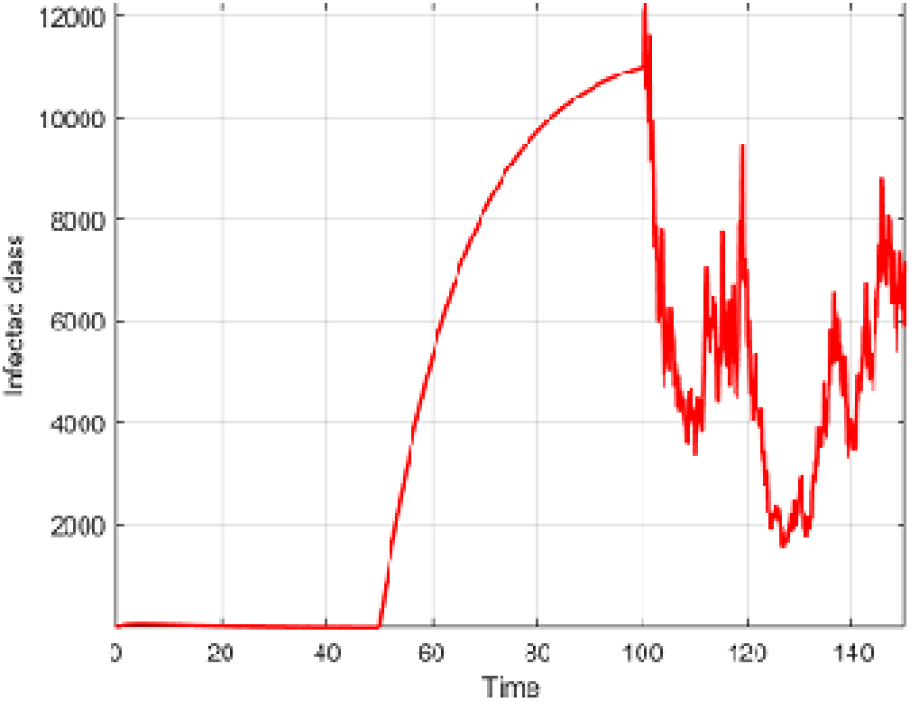
Numerical visualization for infected people for *α* = 1.

**Figure 4.**
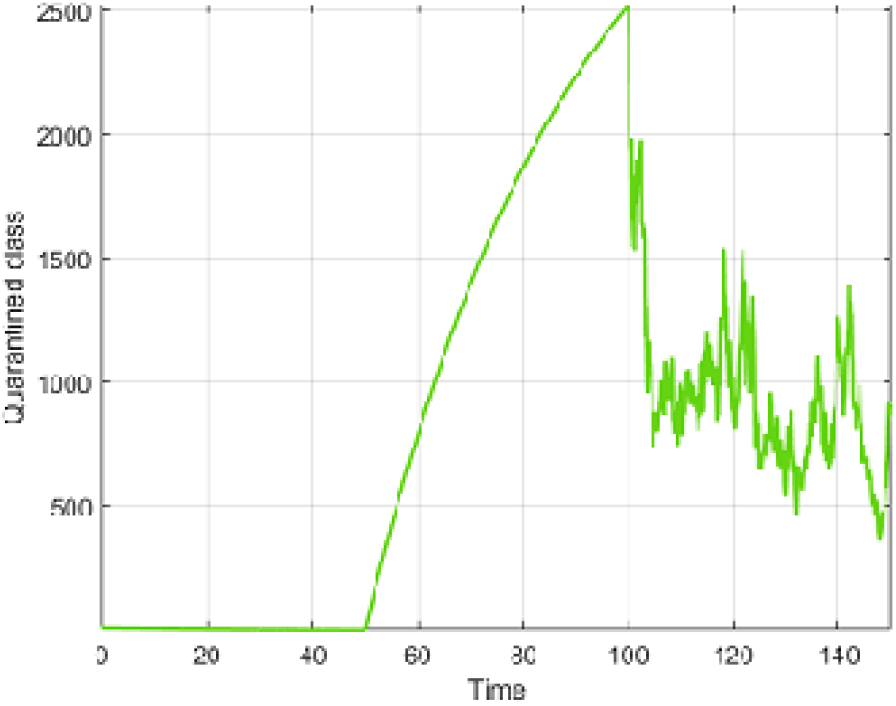
Numerical visualization for quarantined people for *α* = 1.

**Figure 5.**
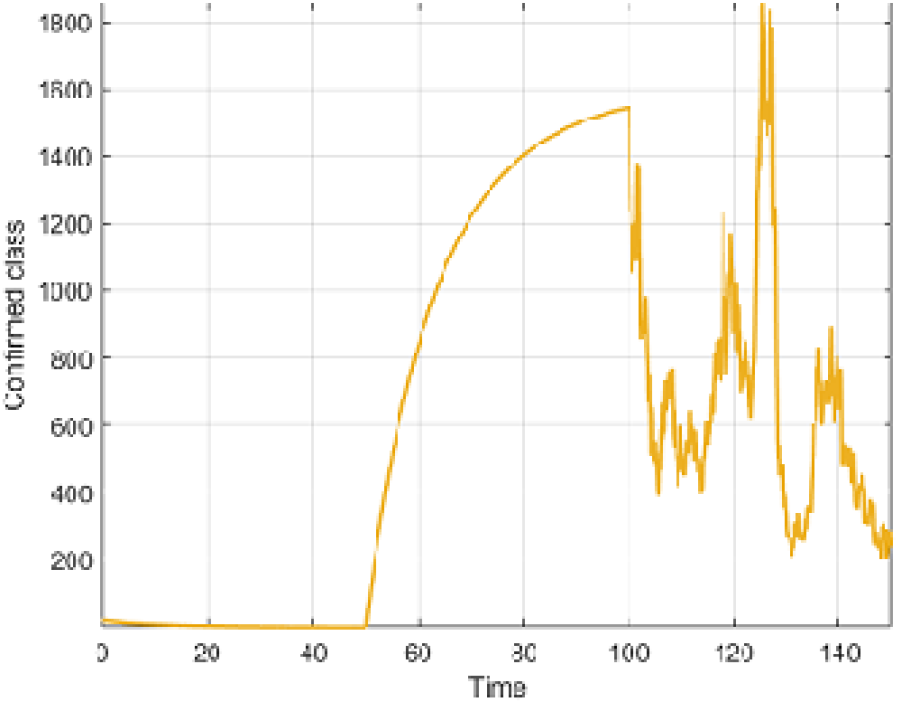
Numerical visualization for confirmed people for *α* = 1.

**Figure 6.**
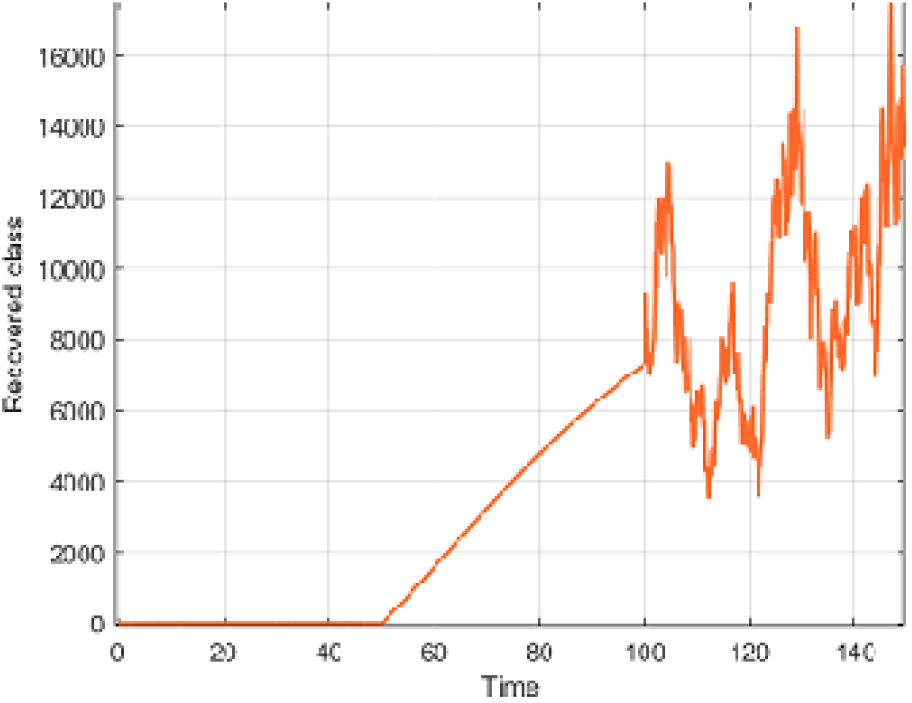
Numerical visualization for recovered people for *α*= 1.

**Figure 7.**
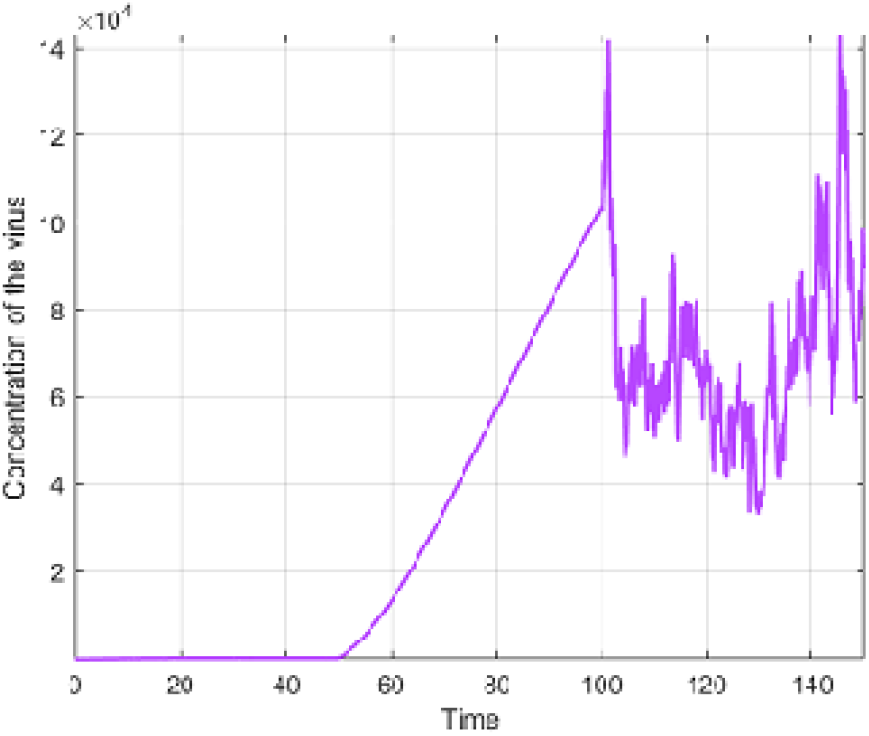
Numerical visualization for concentration of the virus for *α* = 1.

### 6.2 Numerical simulation of Covid-19 model for Case 2

In case two, we consider a given community where the spread displays three processes, including classical behaviors, fading memory behavior and finally stochastic behaviors. In this case, if we consider *T* as the last time of the spread, this is to say, the last day where a new infection occur, then, for the first period of time ranging from 0 to *T*_1_, the mathematical model will be constructed with classical differential operator, the second phrase, the model will be with the Caputo-Fabrizio differential operator and finally stochastic approach will be used at the last phase. The mathematical model explaining this dynamic is then presented below.

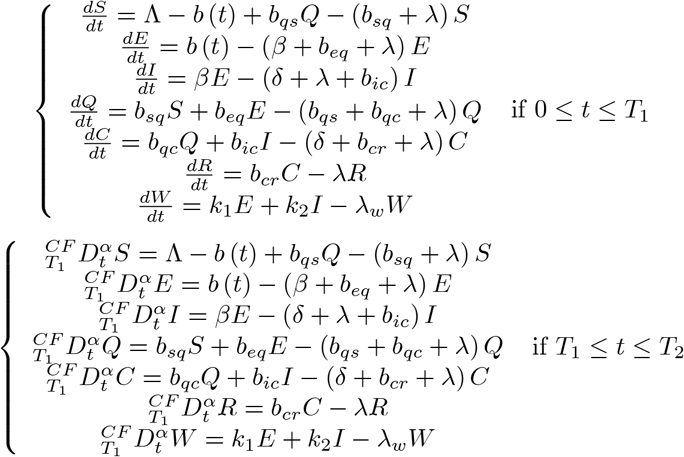

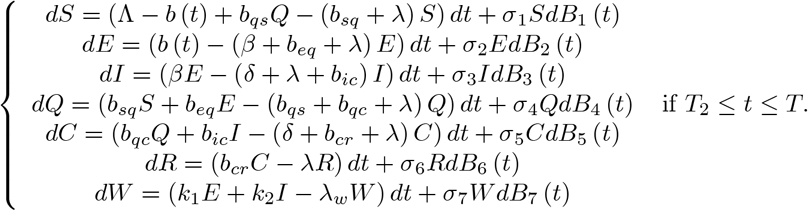

The initial conditions are as follows:

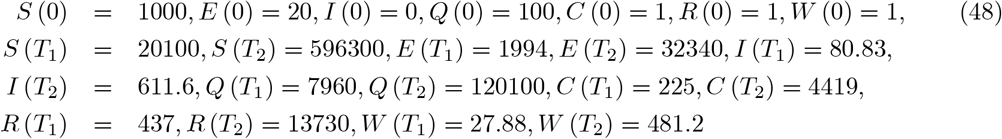

and the parameters are

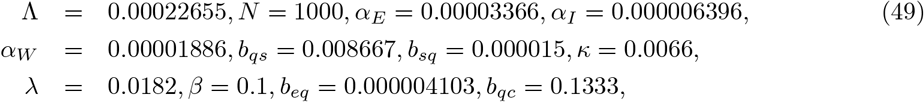

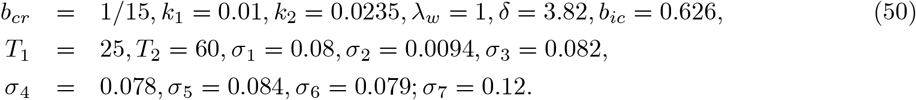

The numerical simulations of model with piecewise differential and integral operators are performed in Figure 8-14.

**Figure 8.**
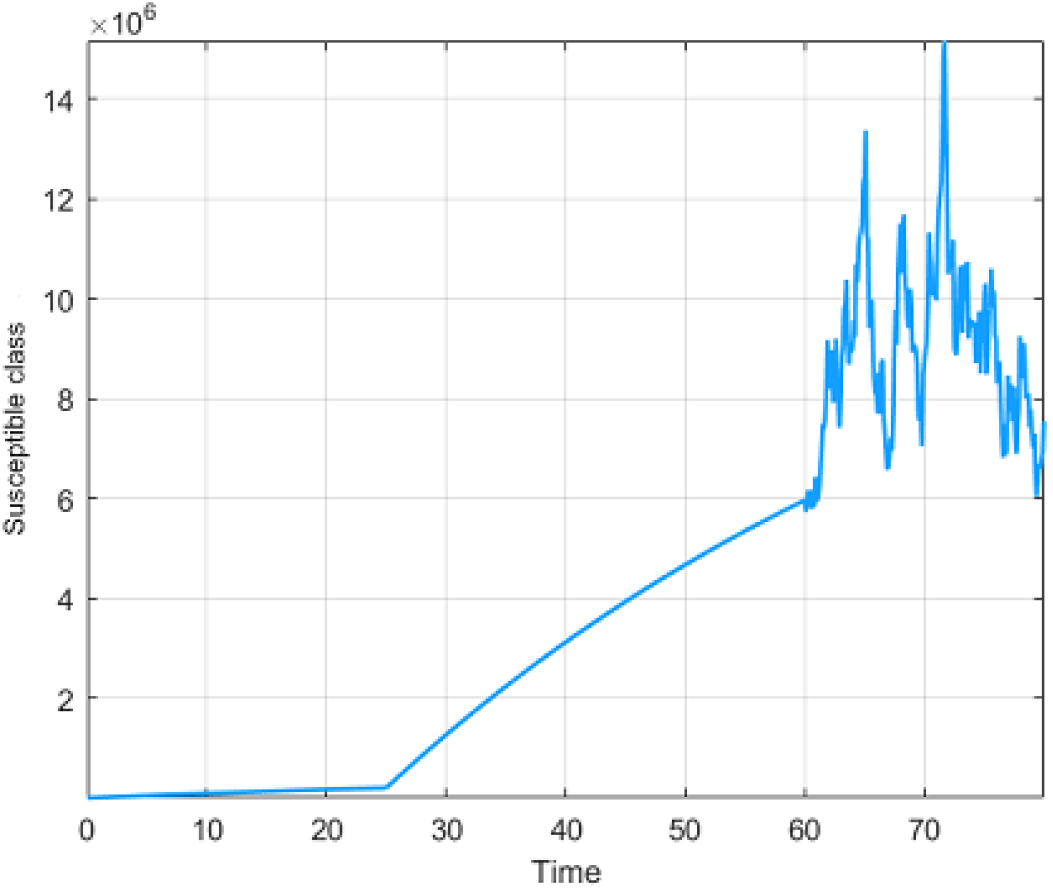
Numerical visualization for susceptible people for *α* = 1.

**Figure 9.**
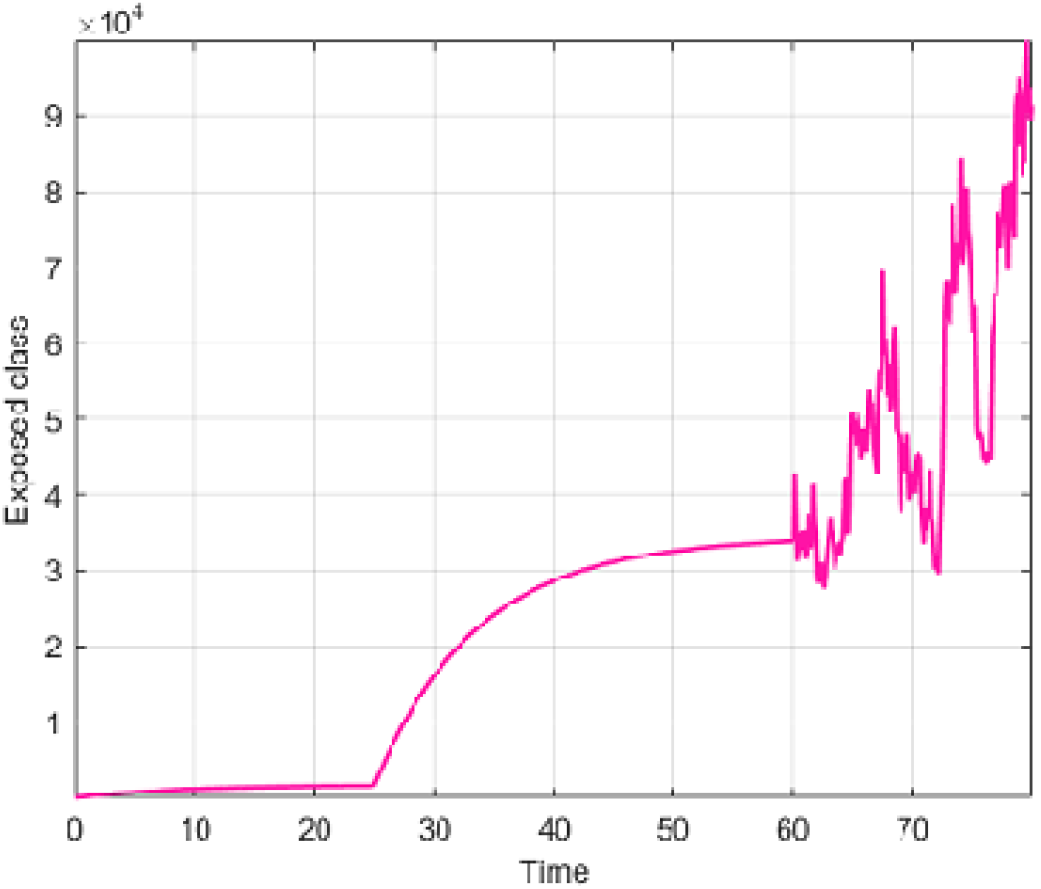
Numerical visualization for exposed people for *α* = 1.

**Figure 10.**
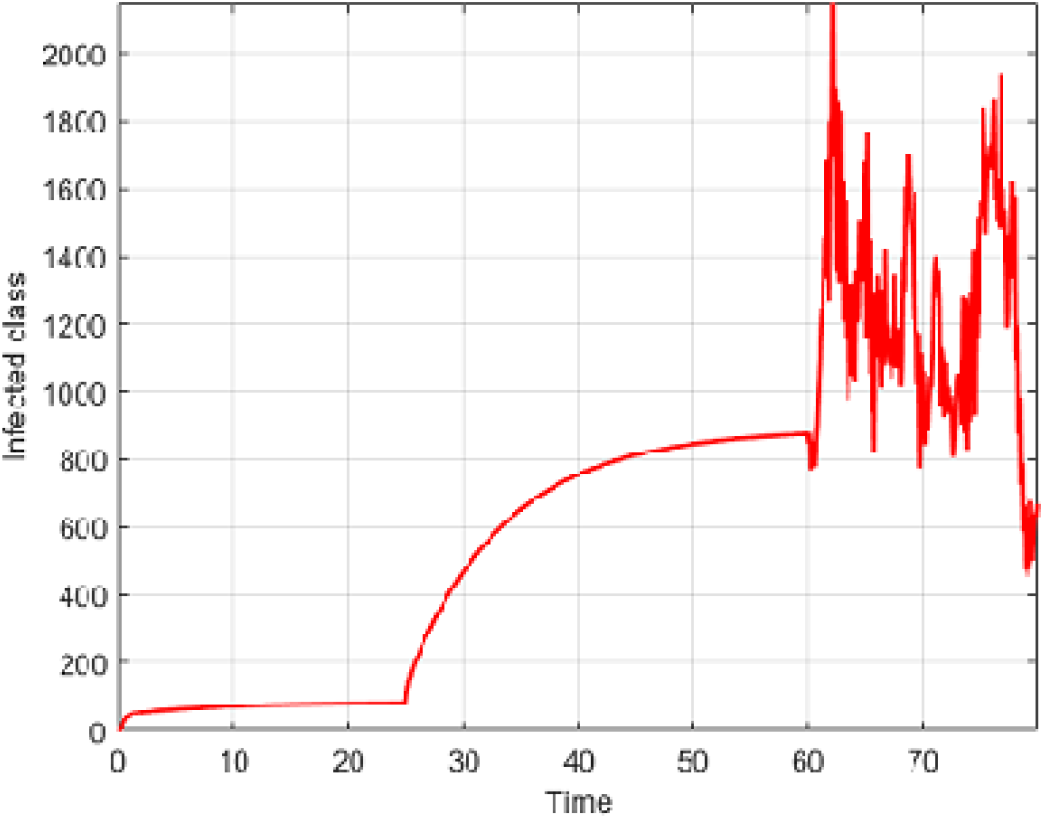
Numerical visualization for infected people for *α*= 1.

**Figure 11.**
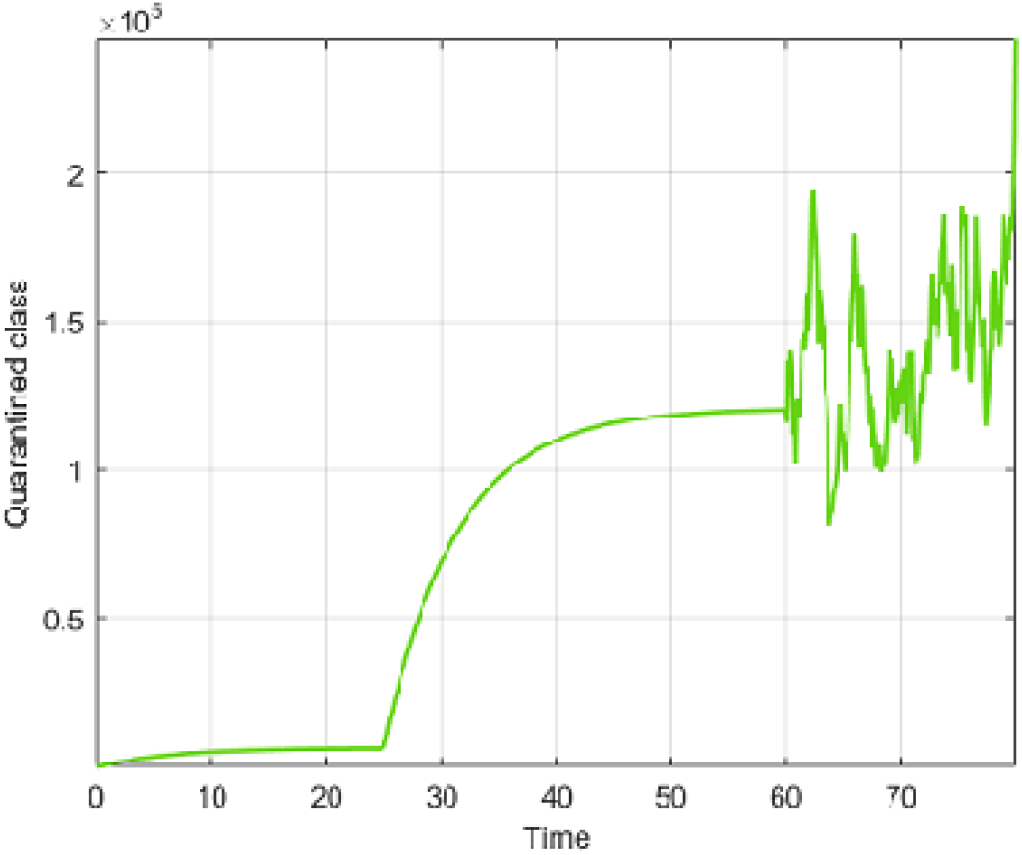
Numerical visualization for quarantined people for *α* = 1.

**Figure 12.**
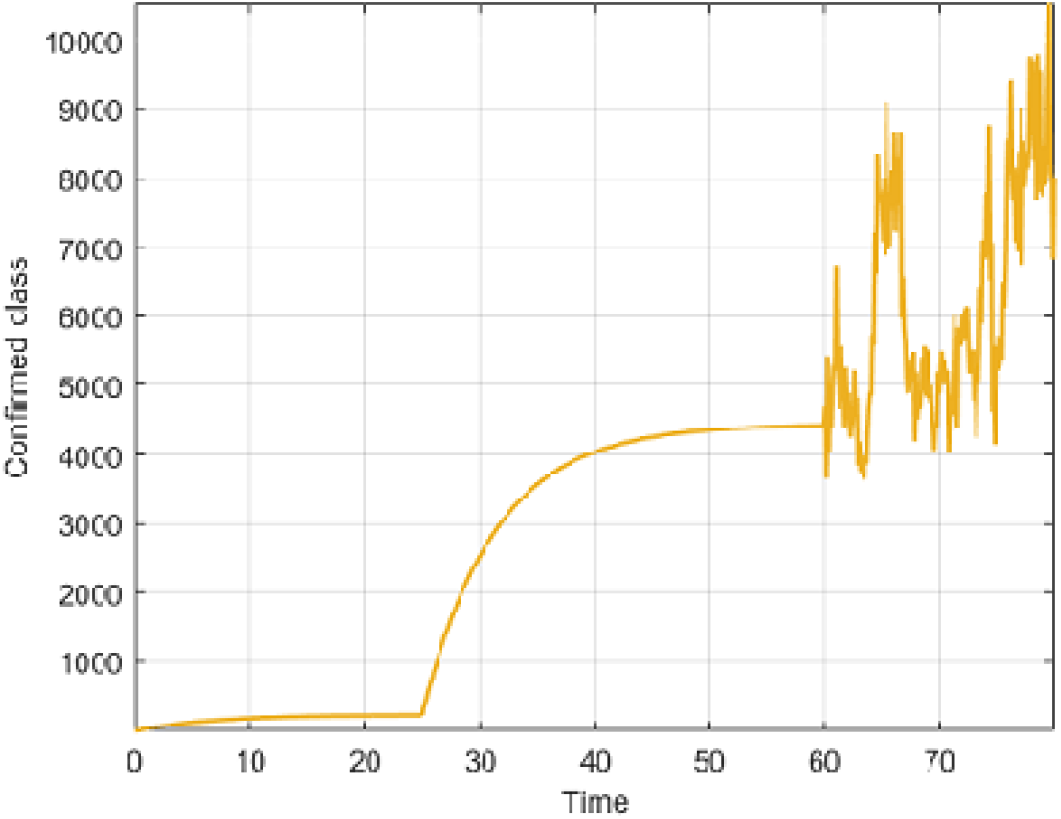
Numerical visualization for confirmed people for *α* = 1.

**Figure 13.**
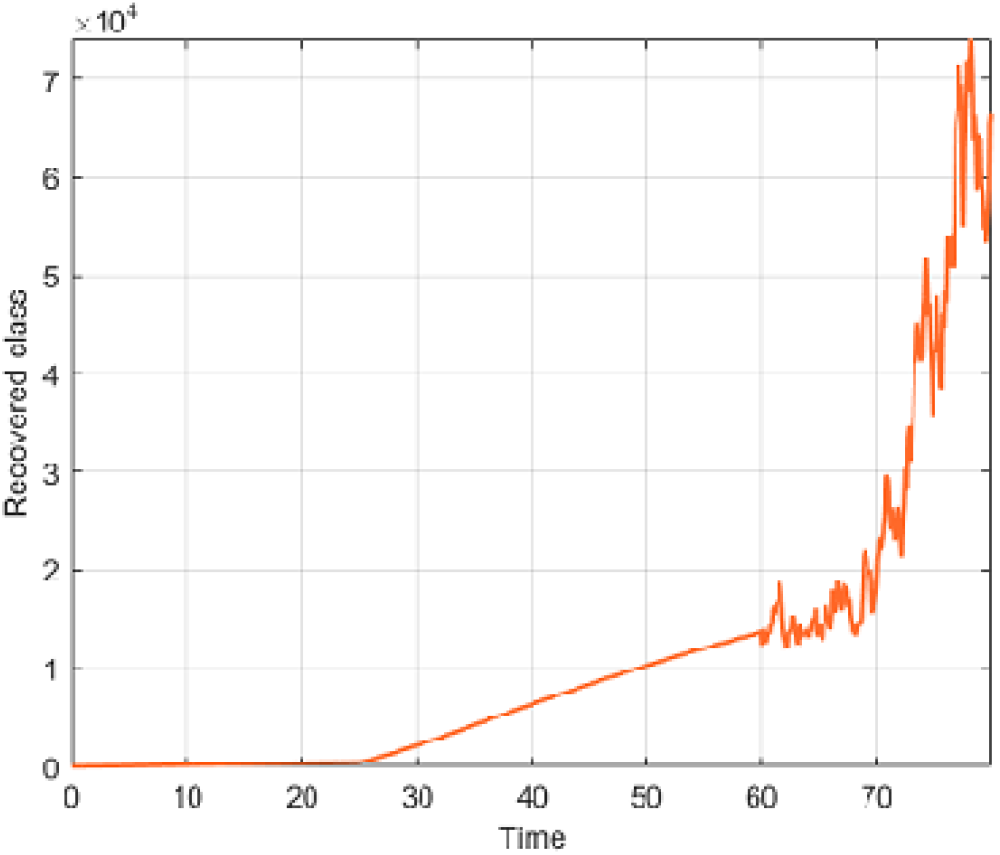
Numerical visualization for recovered people for *α* = 1.

**Figure 14.**
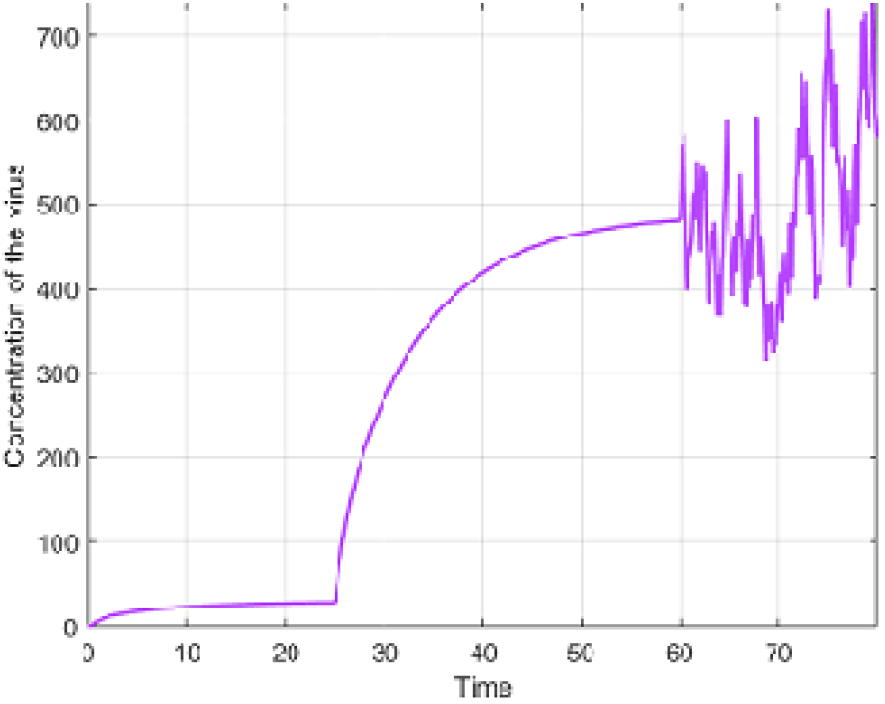
Numerical visualization for concentration of the virus for *α* = 1.

In Figure 15-21, the numerical simulations of model with piecewise derivative are depicted with

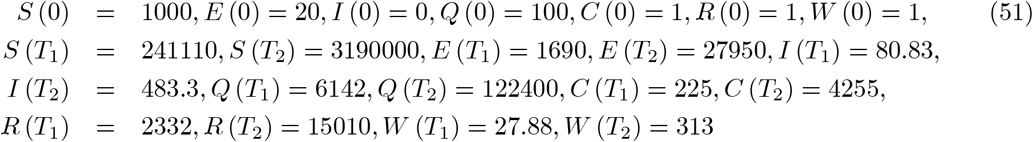

and the parameters

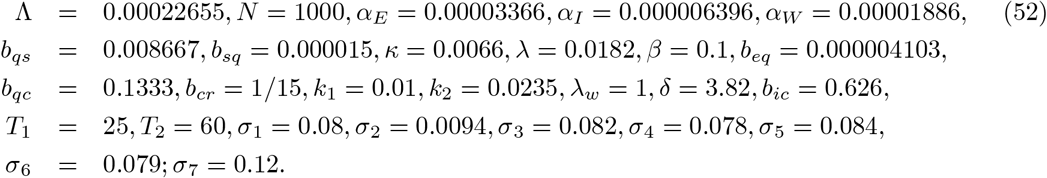

**Figure 15.**
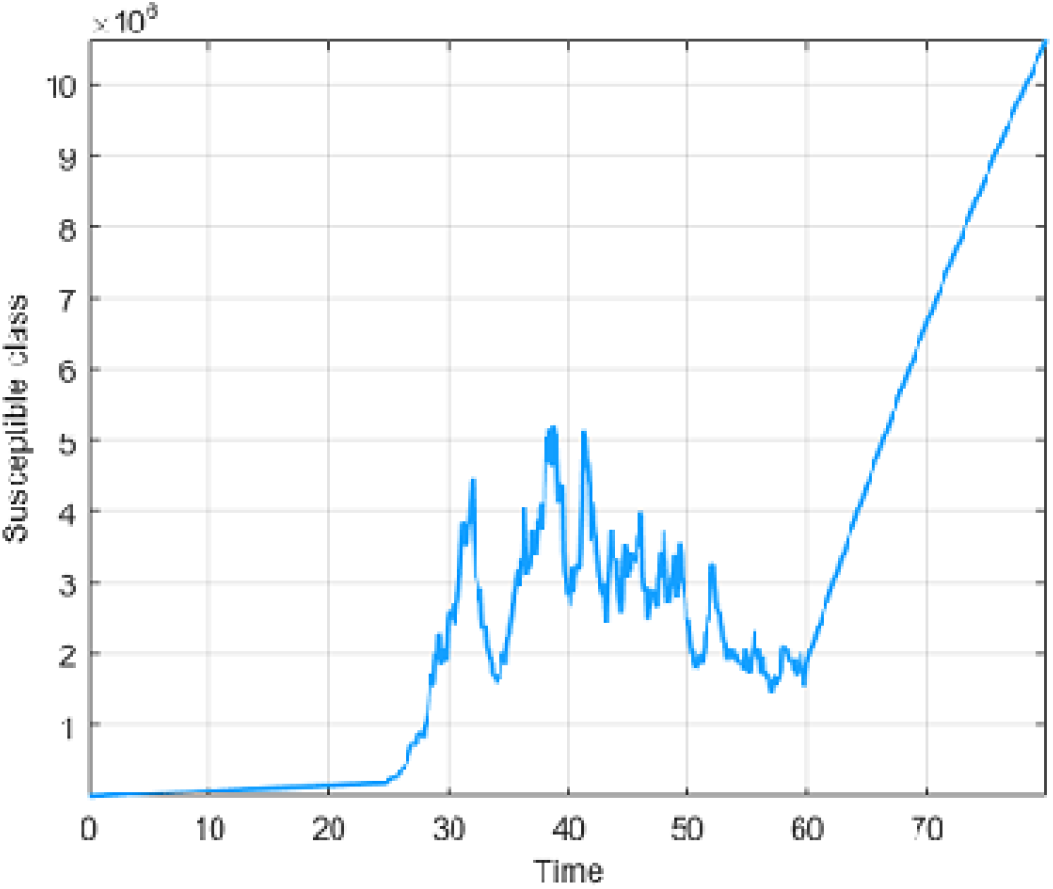
Numerical visualization for susceptible people for *α* = 0.95.

**Figure 16.**
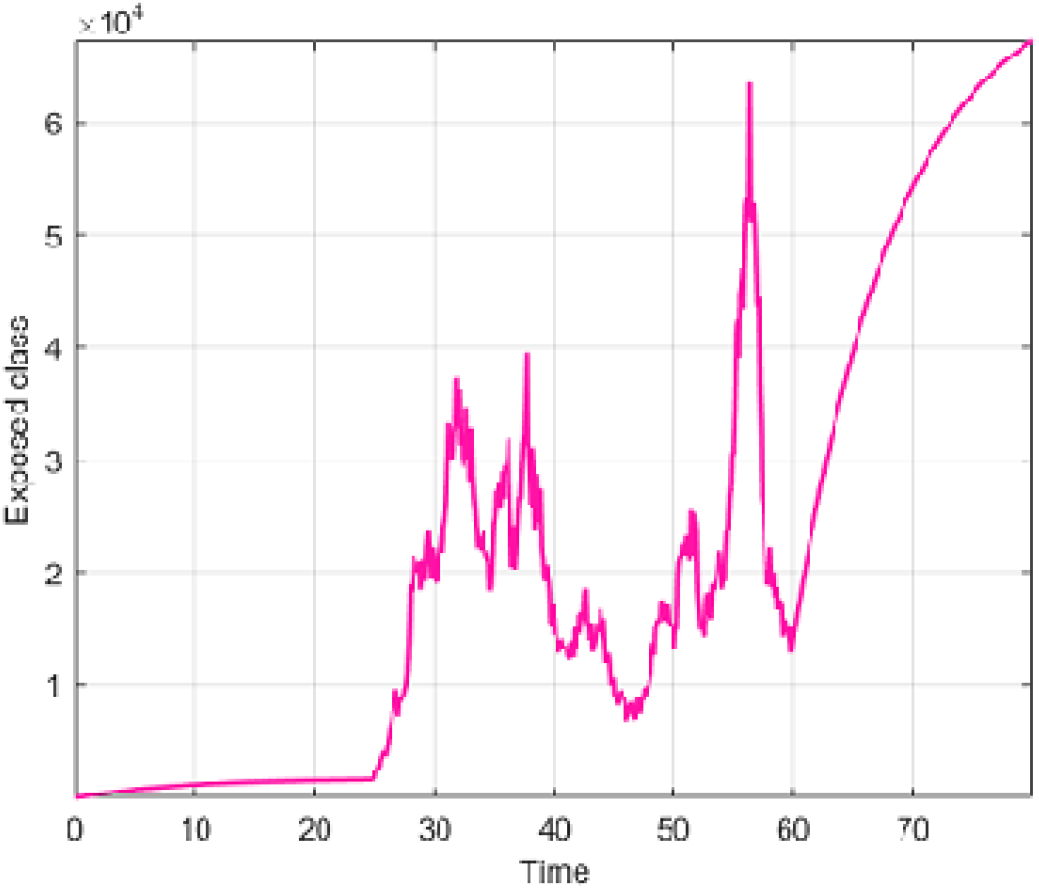
Numerical visualization for exposed people for *α* = 0.95.

**Figure 17.**
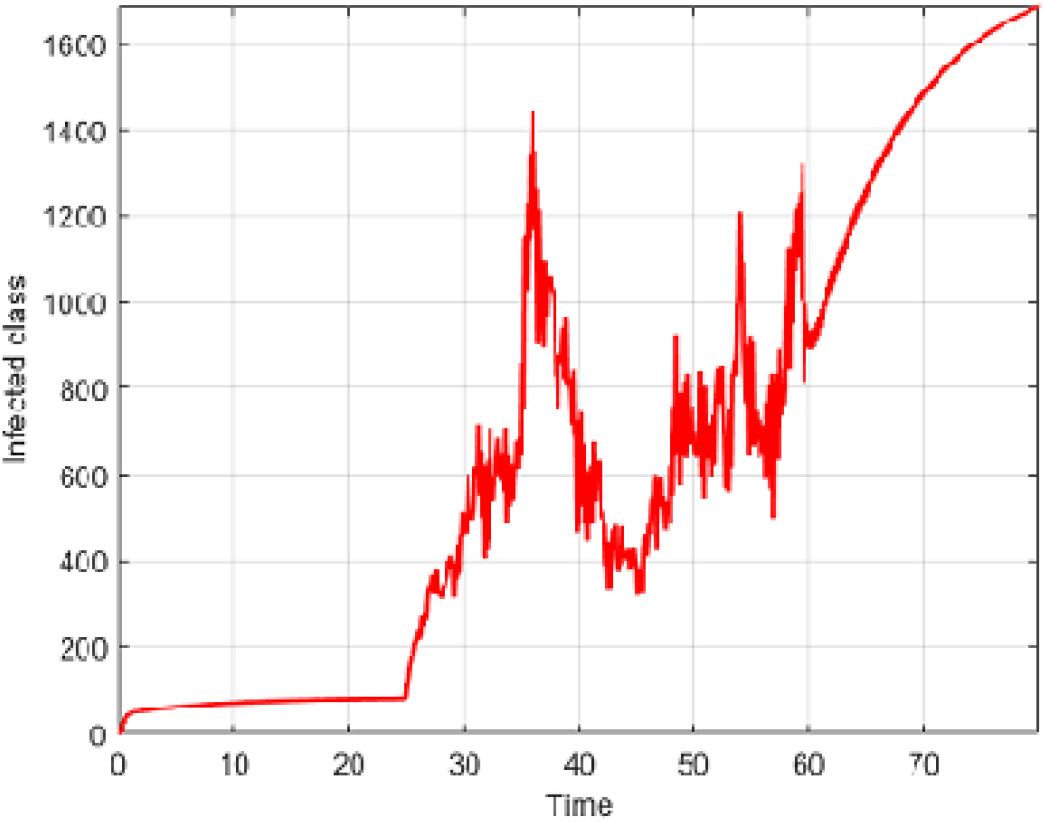
Numerical visualization for infected people for *α* = 0.95.

**Figure 18.**
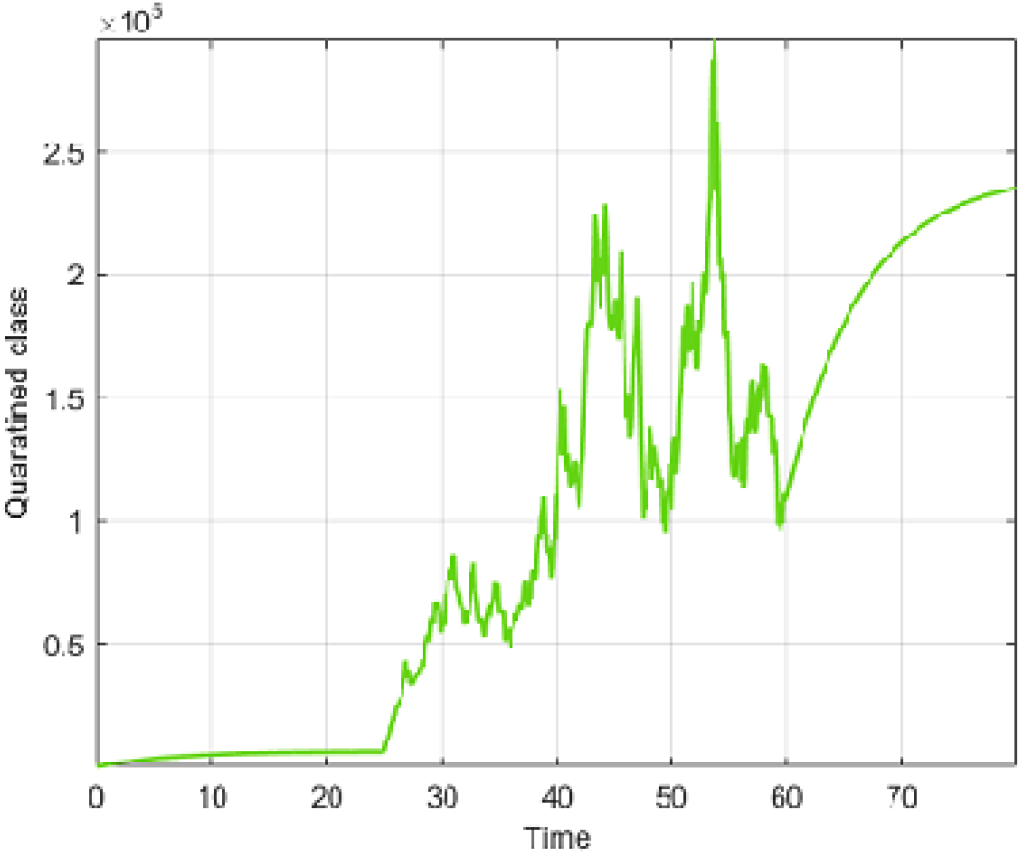
Numerical visualization for quarantined people for *α* = 0.95.

**Figure 19.**
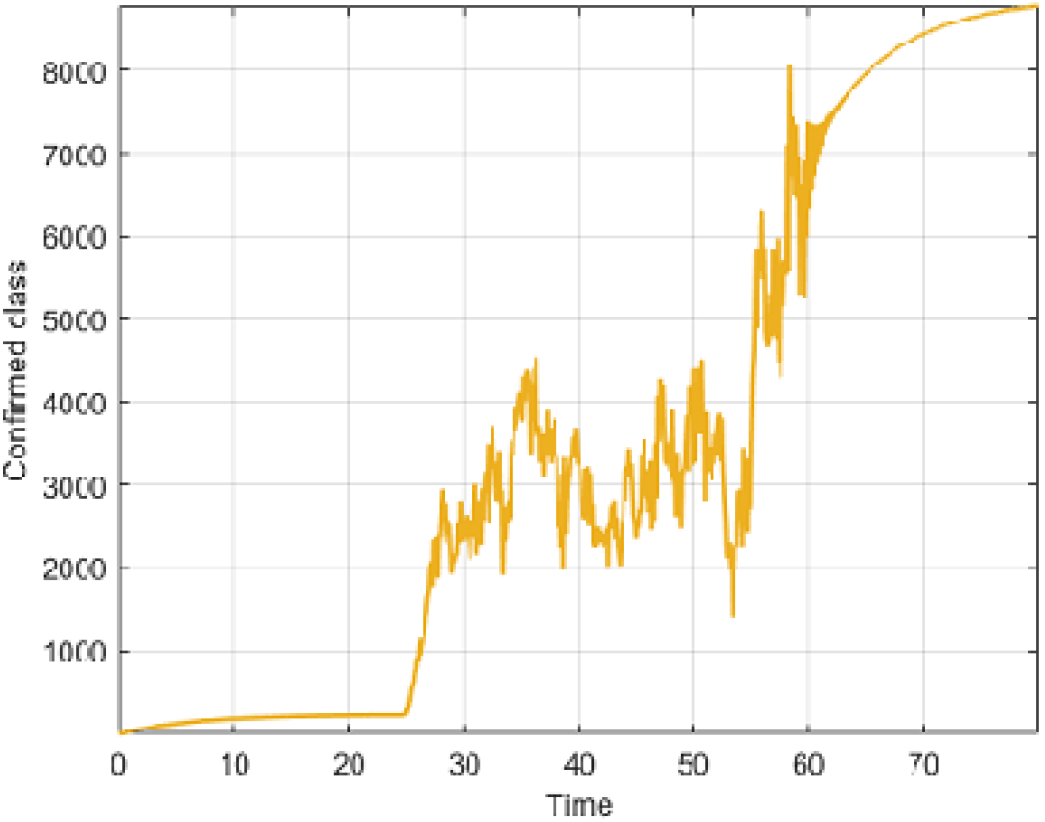
Numerical visualization for confirmed people for *α* = 0.95.

**Figure 20.**
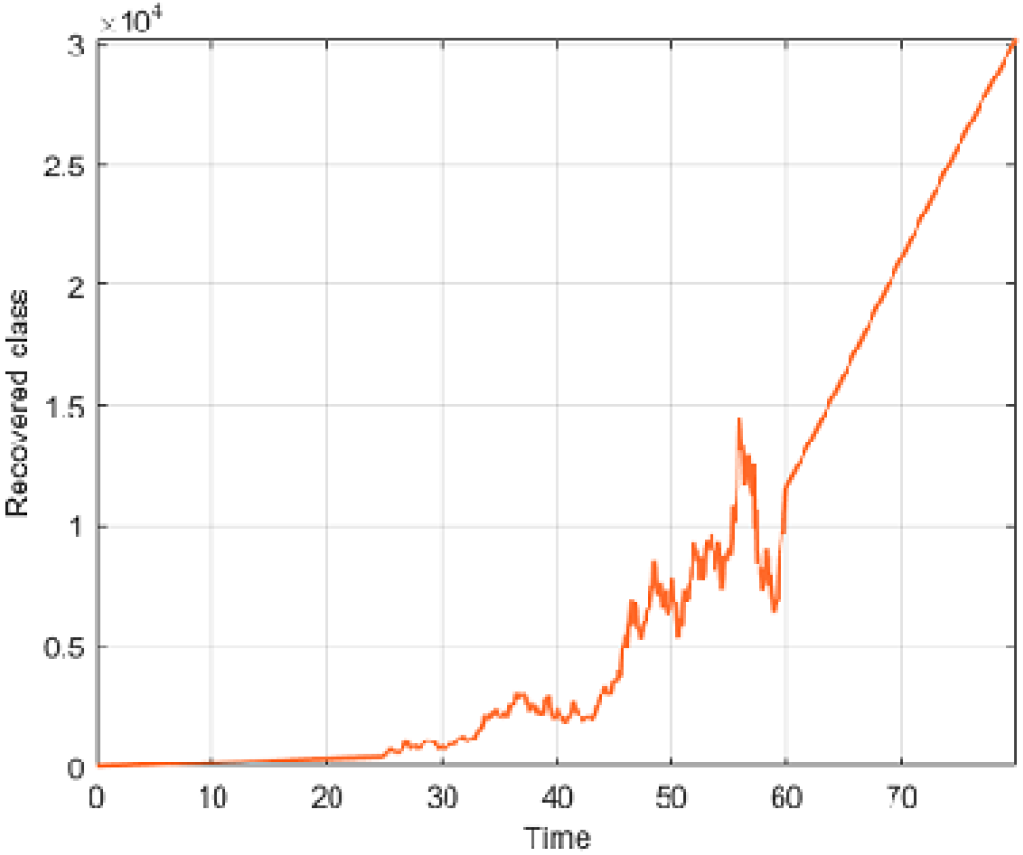
Numerical visualization for recovered people for *α*= 0.95.

**Figure 21.**
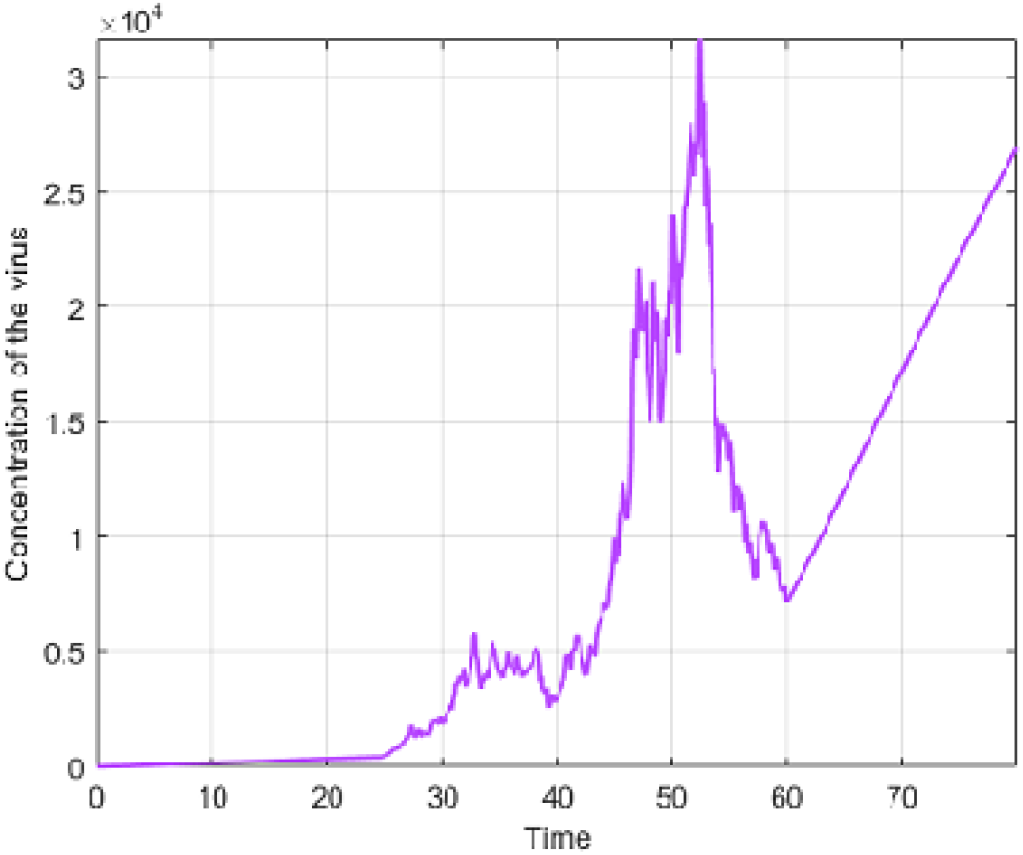
Numerical visualization for concentration of the virus for *α* = 0.95.

### 6.3 Numerical solution of Covid-19 model for Case 3

In case three, assuming a given country within which the spread displays three processes, including classical behaviors, a passage from stretched exponential to power law behavior and finally stochastic behaviors. In this case, if we consider *T* as the last time of the spread, this is to say, the last day where a new infection occur, then, for the first period of time ranging from 0 to *T*_1_, the mathematical model will be constructed with classical differential operator, the second phrase, the model will be with the Atangana-Baleanu fractional differential operator and finally stochastic approach will be used at the last phase. The mathematical model explaining this dynamic is then presented below.

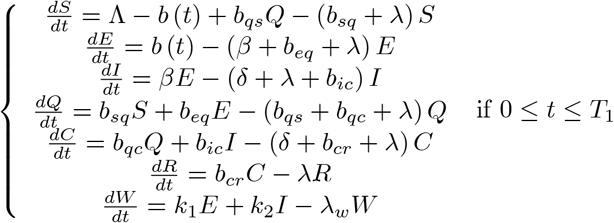

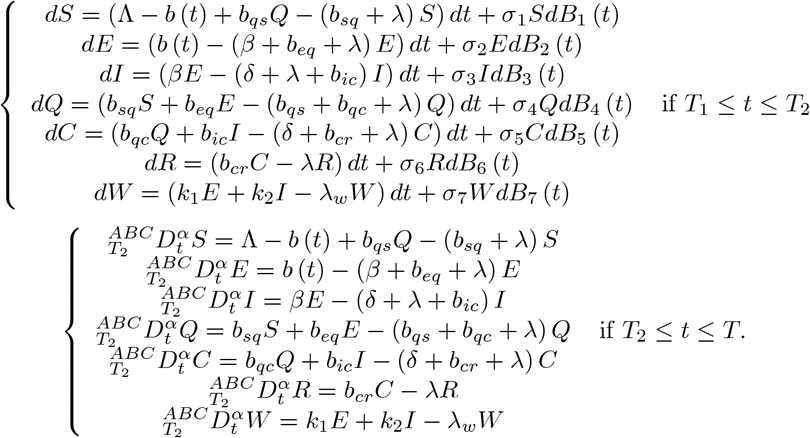

In Figure 22-28, the numerical simulations of model with piecewise derivative are depicted with

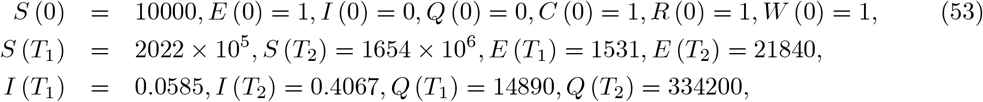

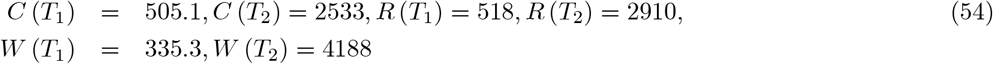

and the parameters

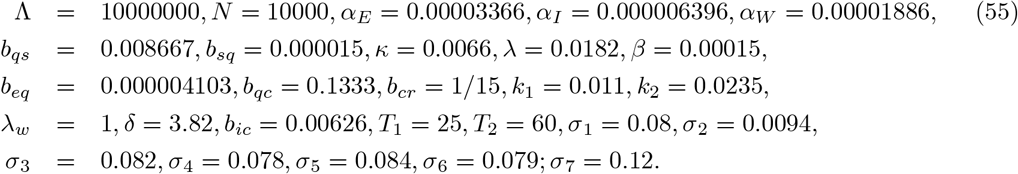

**Figure 22.**
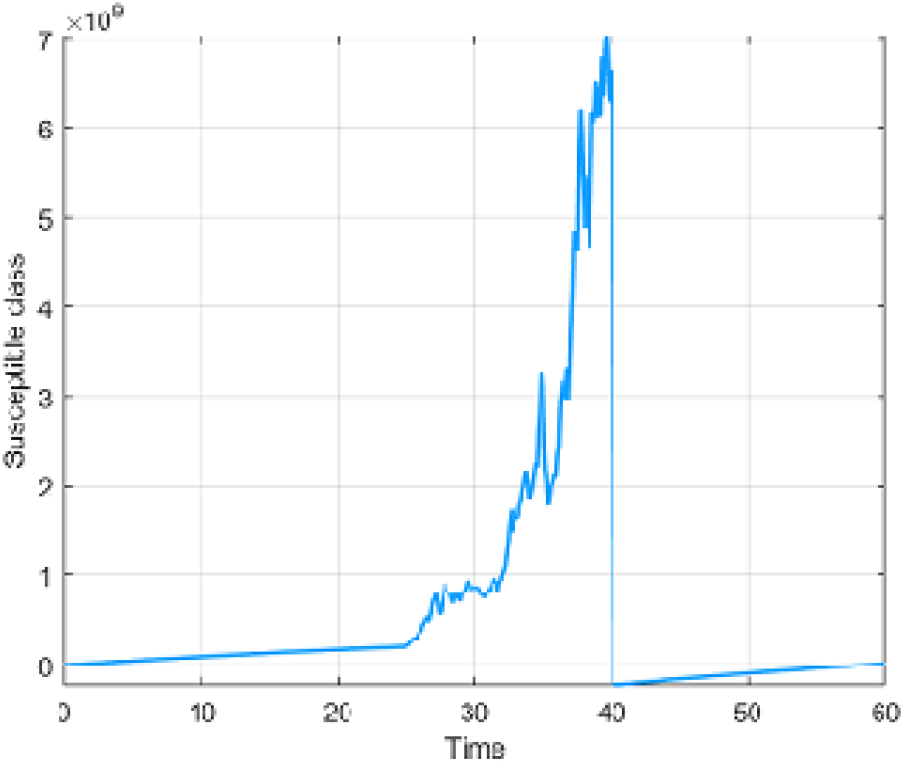
Numerical visualization for susceptible people for *α* = 0.95.

**Figure 23.**
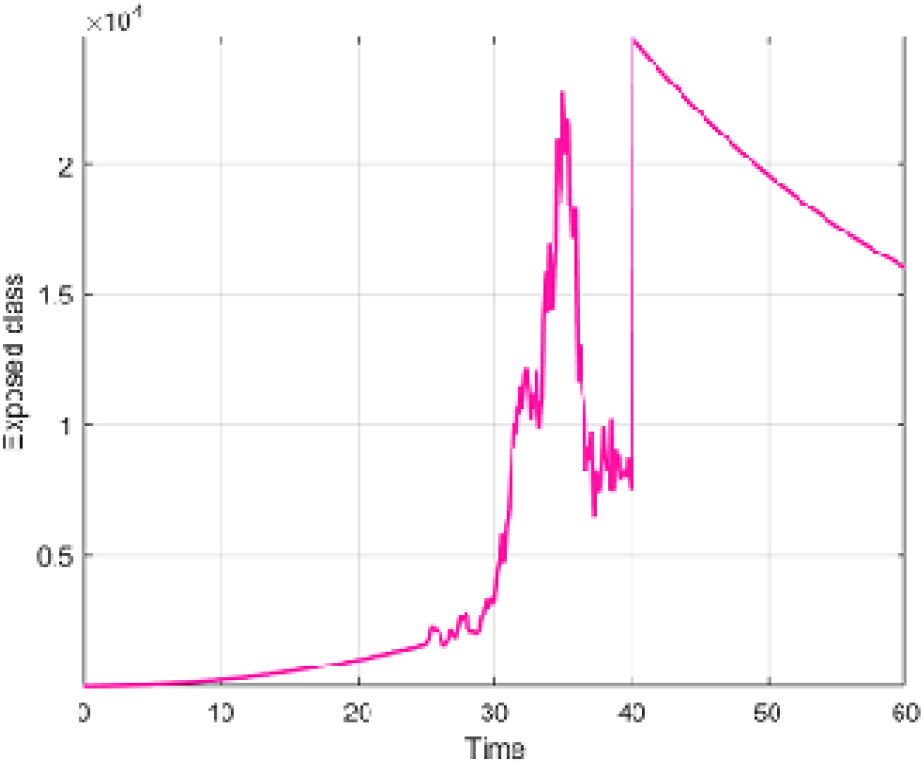
Numerical visualization for exposed people for *α* = 0.95.

**Figure 24.**
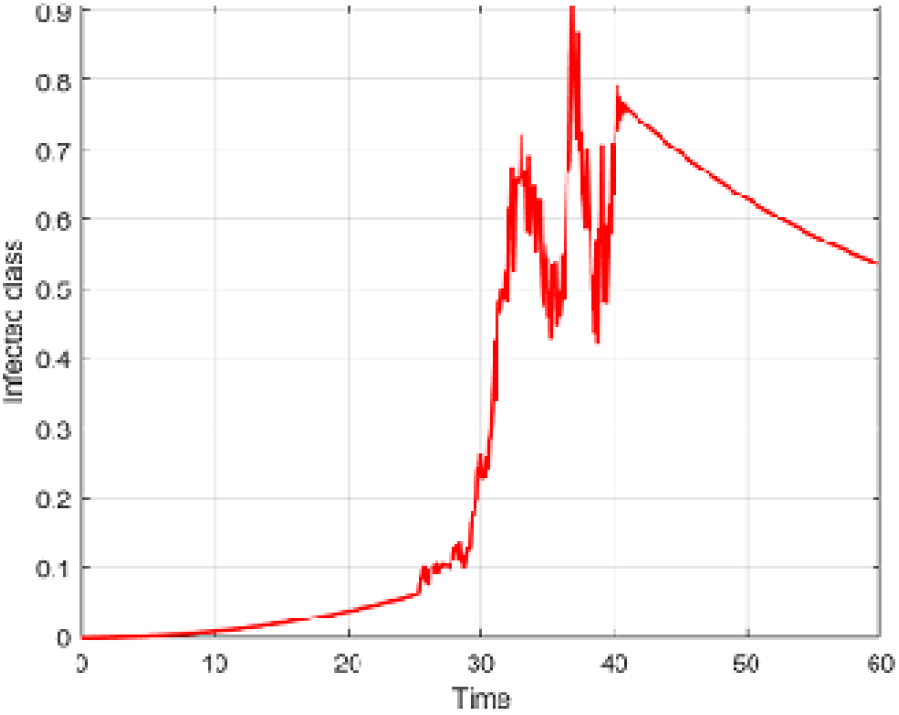
Numerical visualization for infected people for *α*= 0.95.

**Figure 25.**
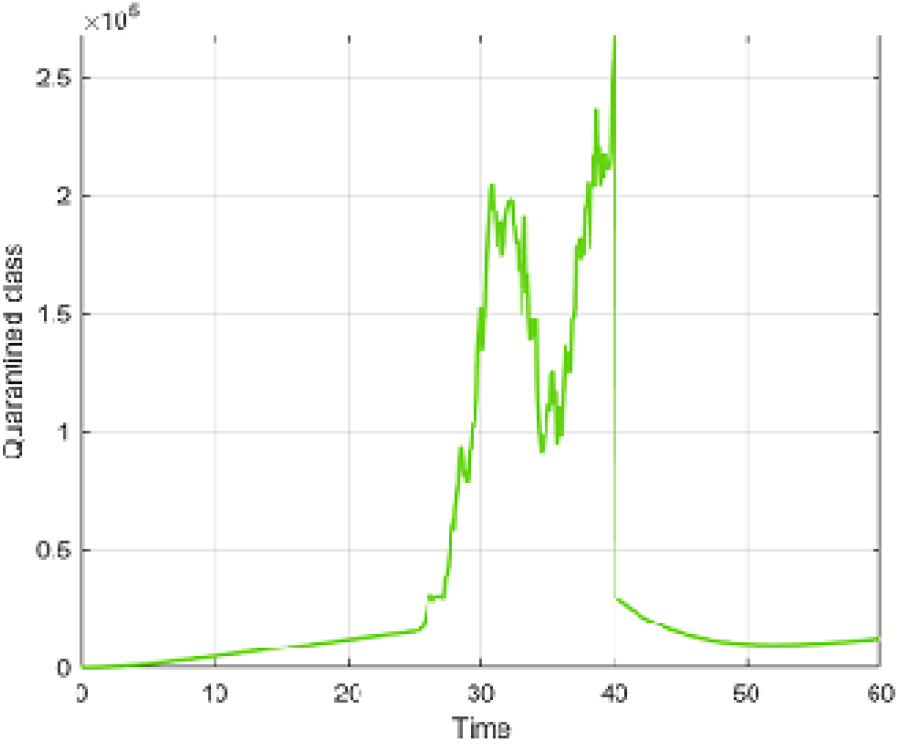
Numerical visualization for quarantined people for *α*= 0.95.

**Figure 26.**
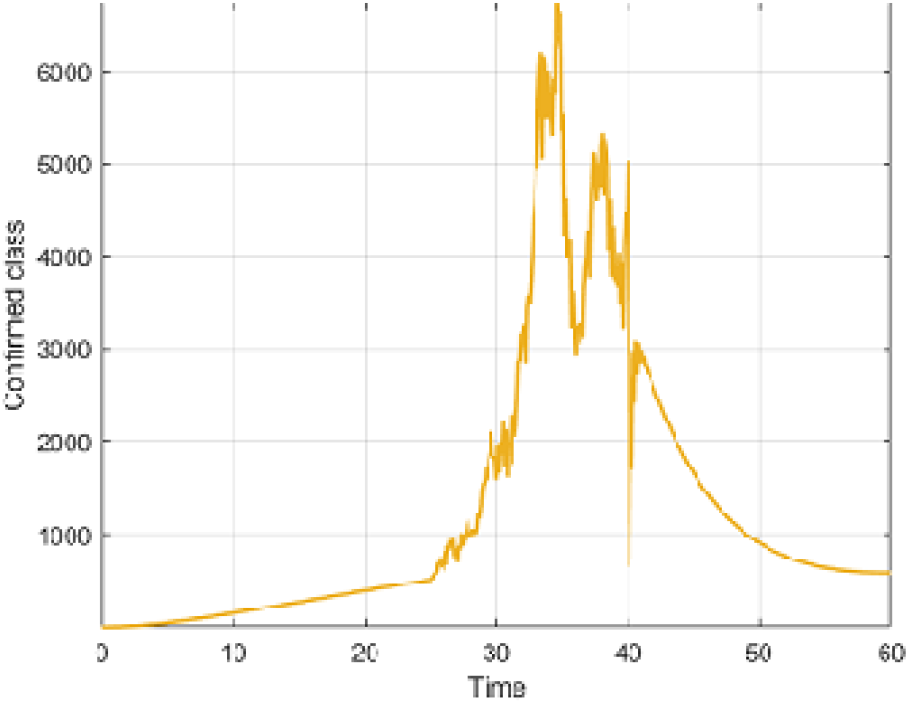
Numerical visualization for confirmed people for *α* = 0.95.

**Figure 27.**
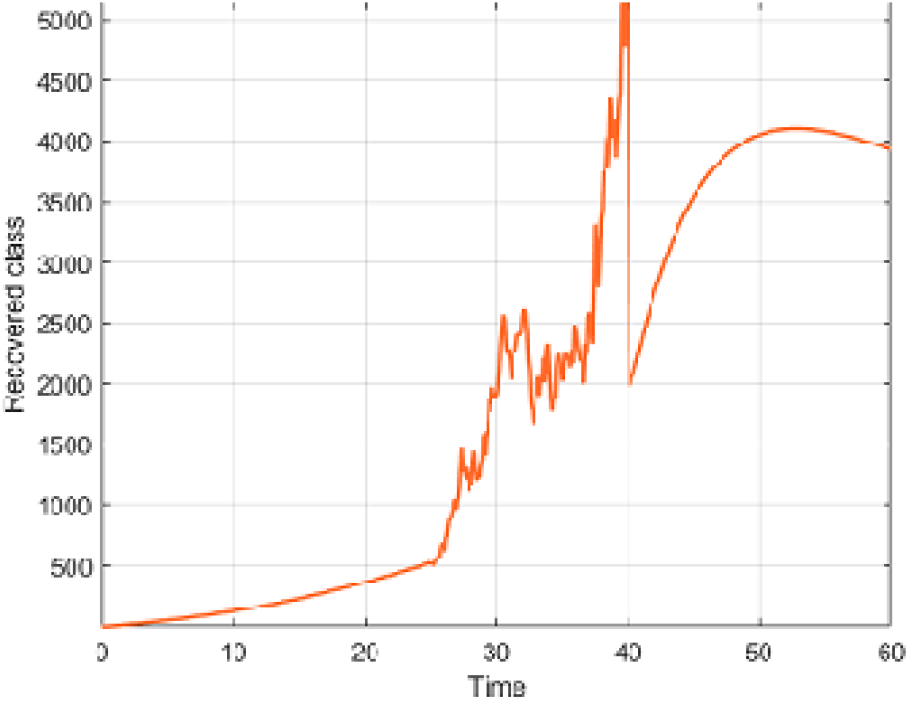
Numerical visualization for recovered people for *α* = 0.95.

**Figure 28.**
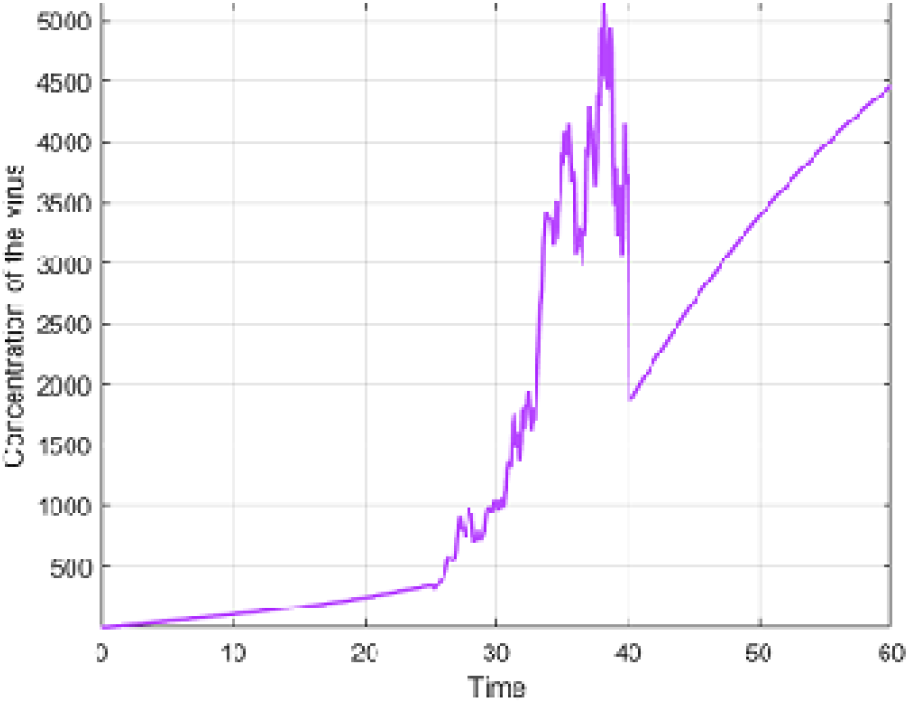
Numerical visualization for concentration of the virus for *α*= 0.95.

## 7 Comparison between piecewise model and real data for some countries

### 7.1 Comparison between a piecewise model and real data in Turkey

In this subsection, we compare modified model by using model that we introduced in the previous chapters with daily number of cases between March 11 and April 20, 2021 in Turkey. Since it will not be easy to predict such long time interval data, to achieve greater compatibility between model that we will suggest and data, we will write our model in more pieces. In other words, although we presented our model in 3 pieces in the previous chapters, of course, we should know that we can write the model in more pieces in order to capture more reality in modeling daily problems. Thus, it will be inevitable for us to achieve our goal. But since this situation brings much more complexity with it, that’s why let us first present some notations to avoid this complexity. We will use the notation *X*_*i*_, *i* = 1, 2, .., 7 such that

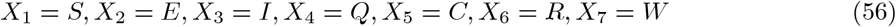

and

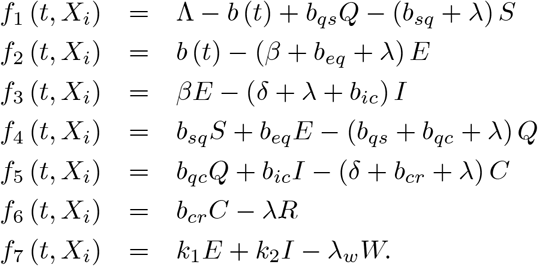

To do comparison, considering above notations, we construct the following model piecewisely

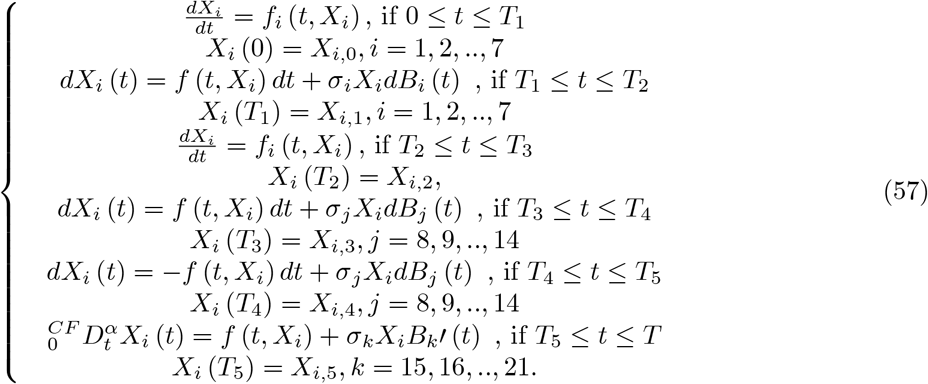

We shall present the parameters and initial conditions that we used for comparison in Turkey. Initial conditions are considered as

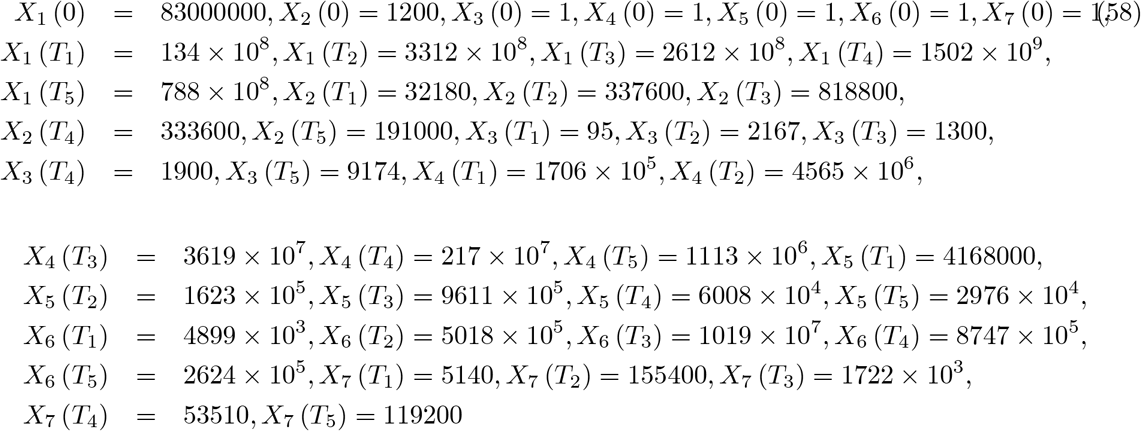

and the parameters are

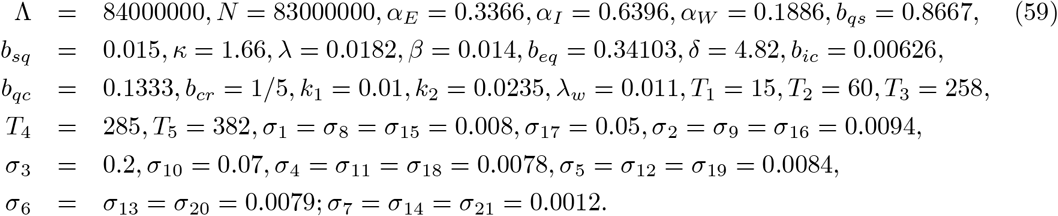

The simulations for comparison are performed using above initial data and parameters in Figure 29 and 30.

**Figure 29.**
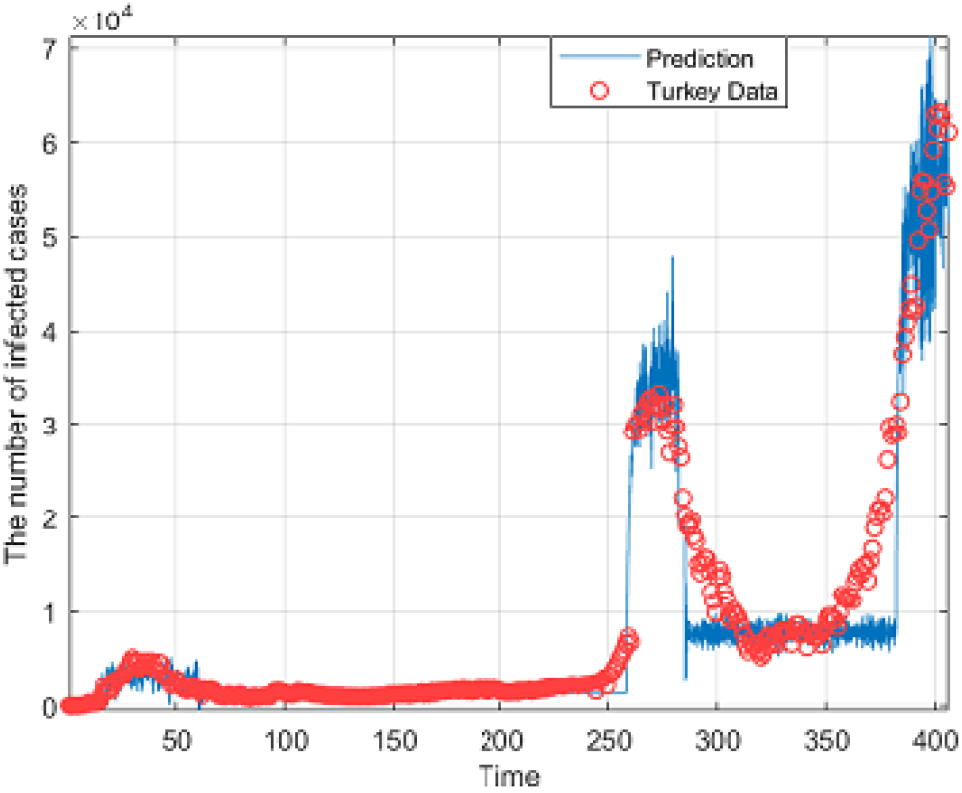
Comparison between piecewise model and real data in Turkey for *α* = 0.98.

**Figure 30.**
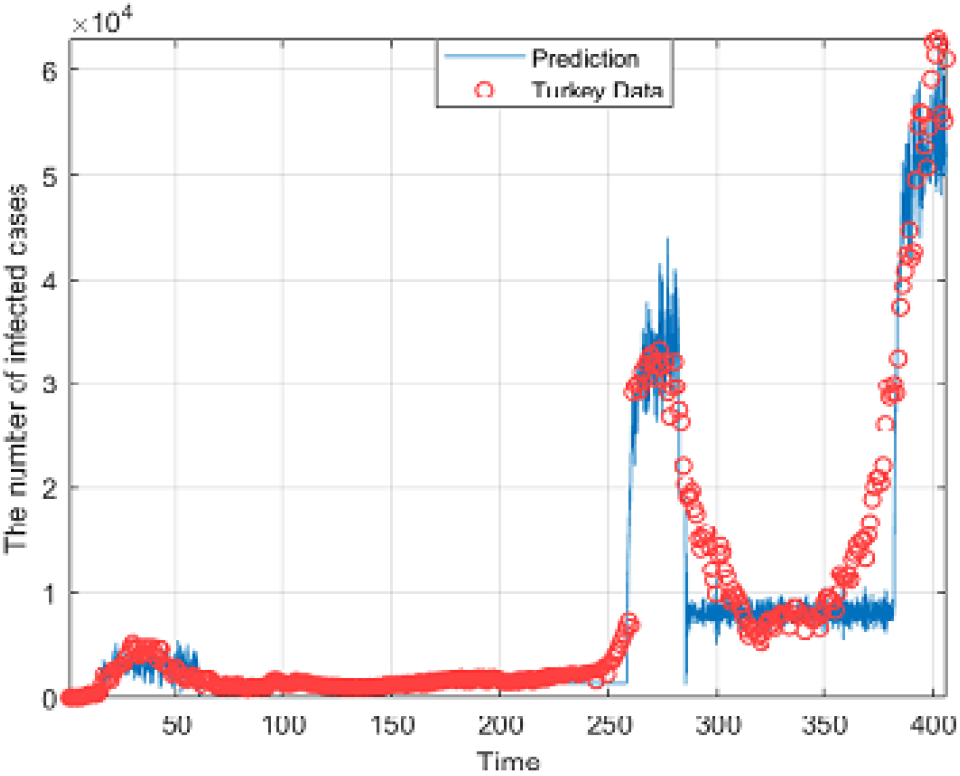
Comparison between piecewise model and real data in Turkey for *α* = 1.

### 7.2 Comparison between piecewise model and real data in Czechia

In this subsection, we compare the daily number of cases between 2 March and 20 April 2021 in Czech Republic with model that will be presented here to analyze the concept we have introduced and to display different processes. Thus, we present the following model which composed of 7 pieces

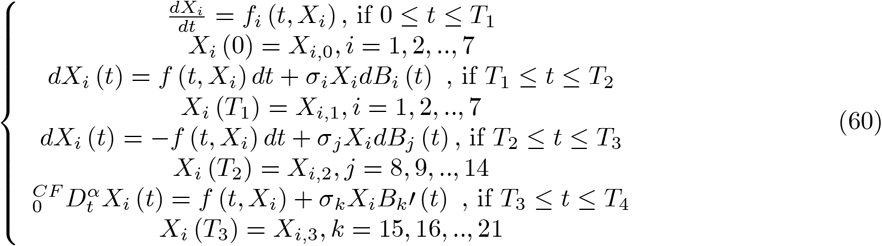

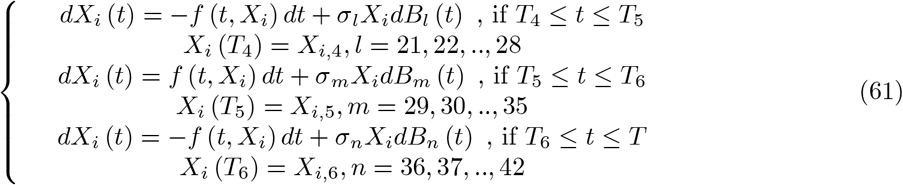

The parameters and initial conditions that we used for comparison in Czechia are given as

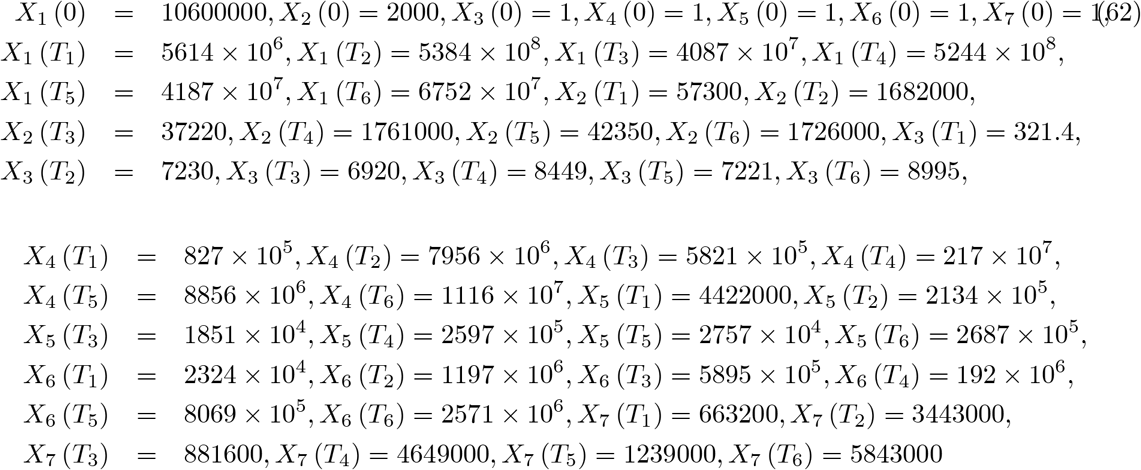

and

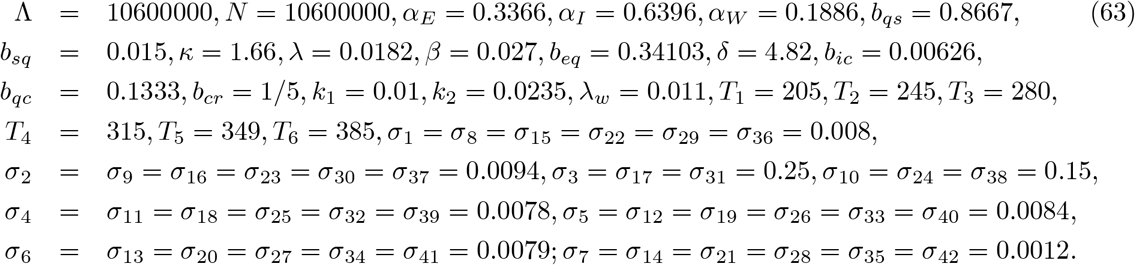

The simulations for comparison are presented using above initial data and parameters in Figure 31 and 32.

**Figure 31.**
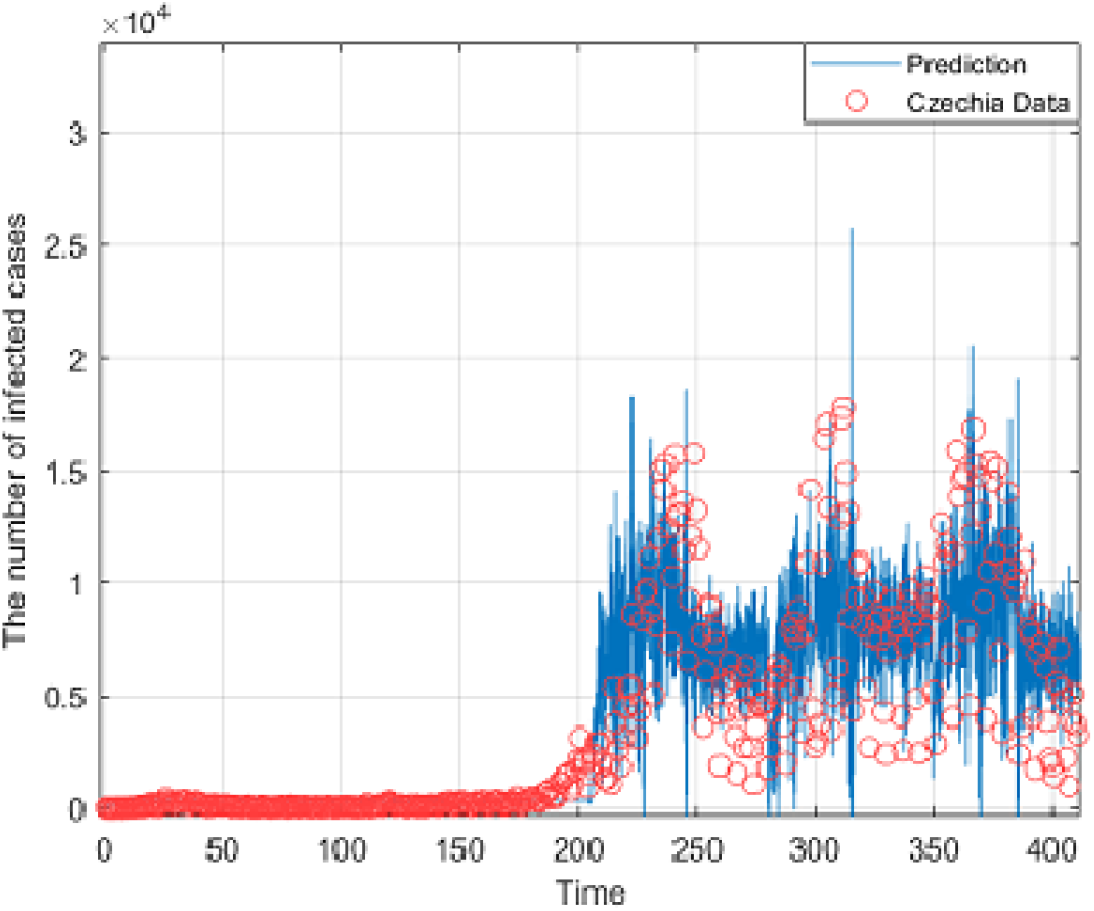
Comparison between piecewise model and real data in Czechia for *α* = 0.985.

**Figure 32.**
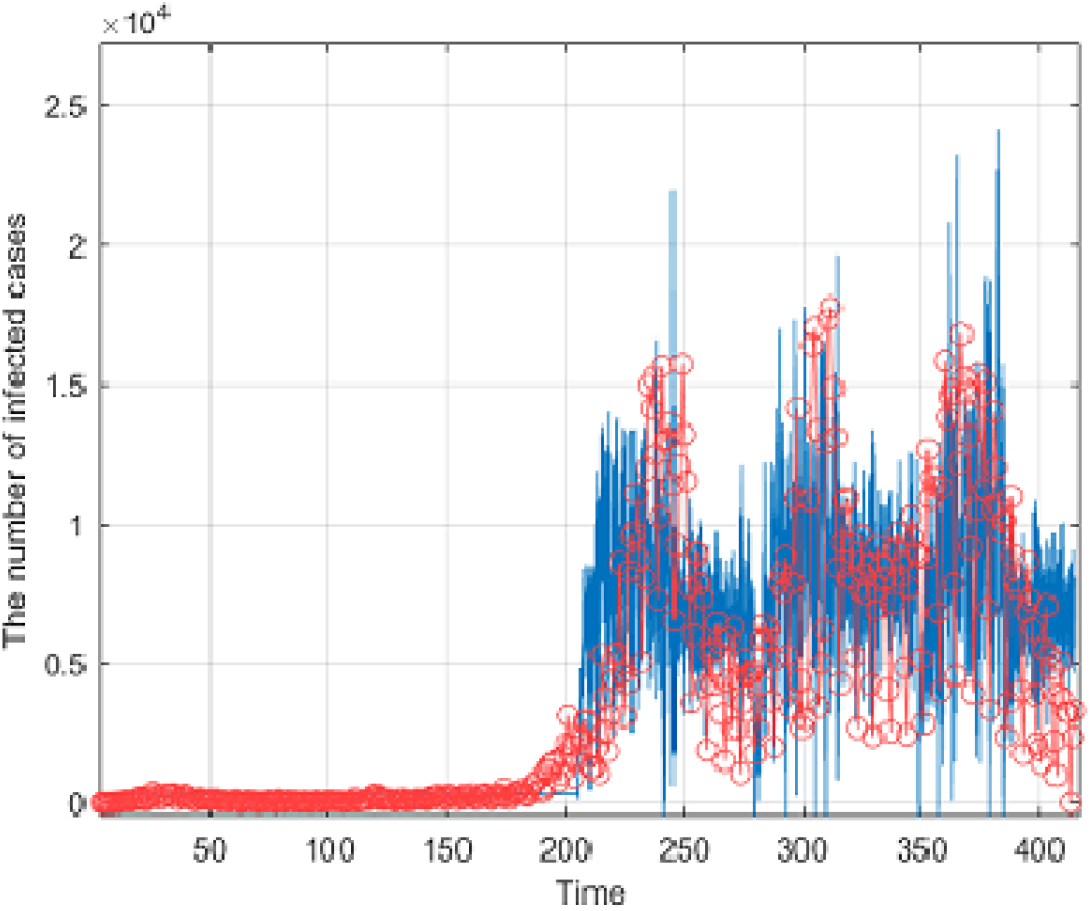
Comparison between piecewise model and real data in Czechia for *α* = 1.

### 7.3 Comparison between piecewise model and real data in Spain

In this subsection, we present a comparison between the daily number of cases on 23 February and 20 April 2021 in Spain and the model that is divided into 9 pieces. To do comparison, considering above notations, we consider the following model

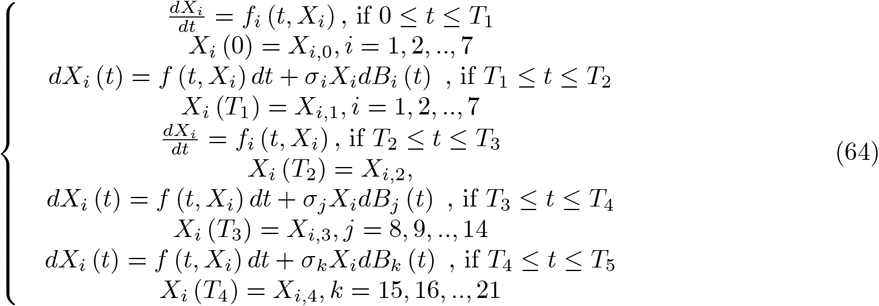

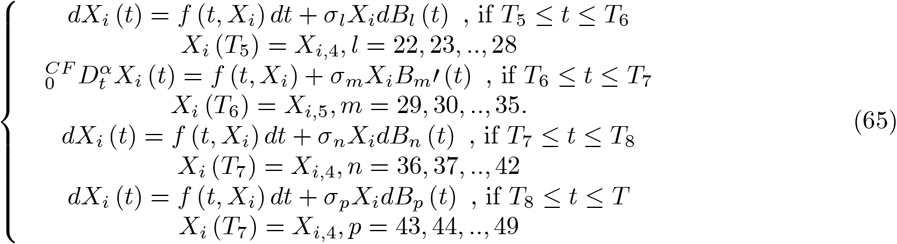

The parameters and initial conditions are as follows

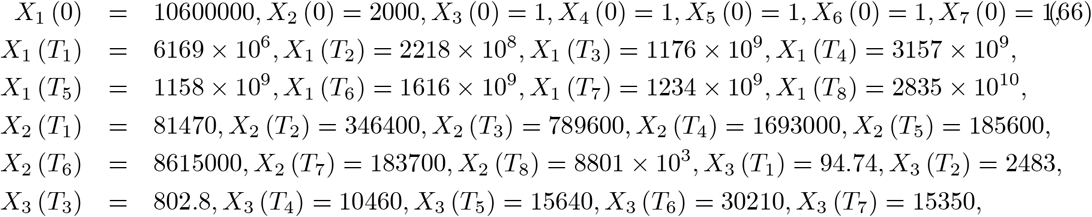

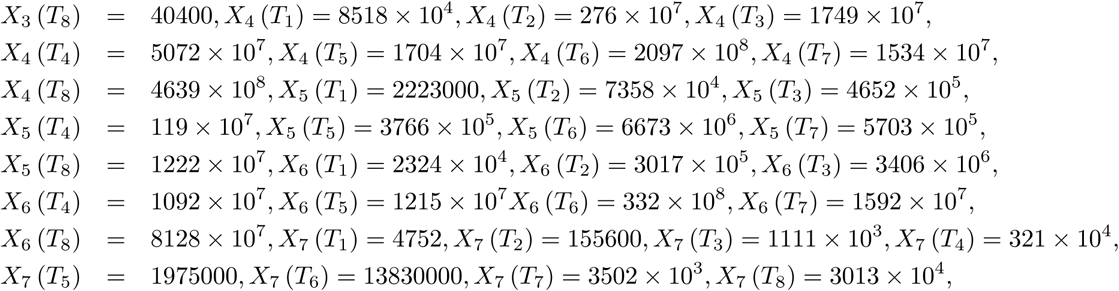

and

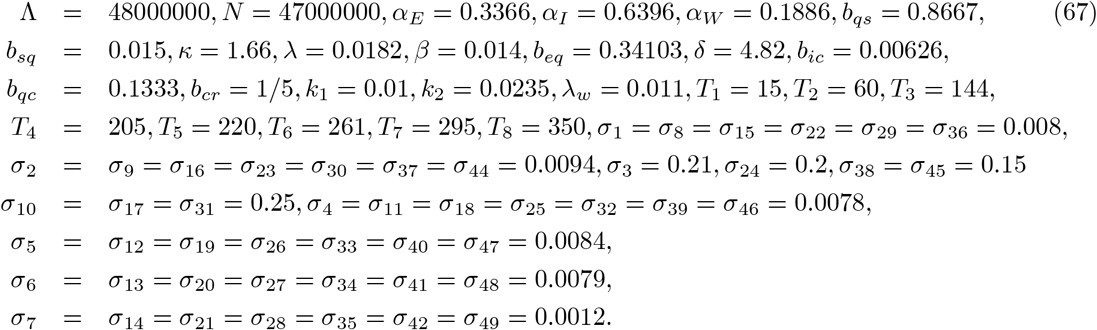

The simulations for comparison are provided using above initial data and parameters in Figure 33 and 34.

**Figure 33.**
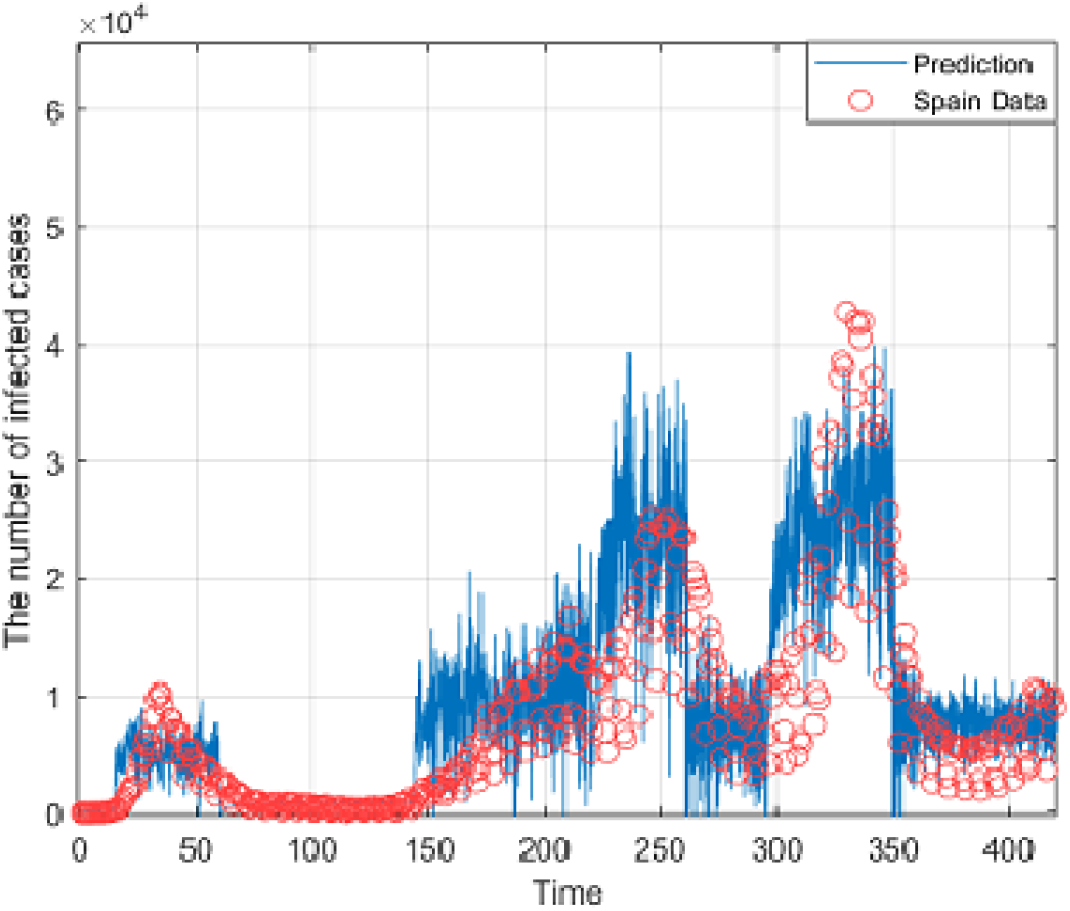
Comparison between piecewise model and real data in Spain for *α* = 0.99.

**Figure 34.**
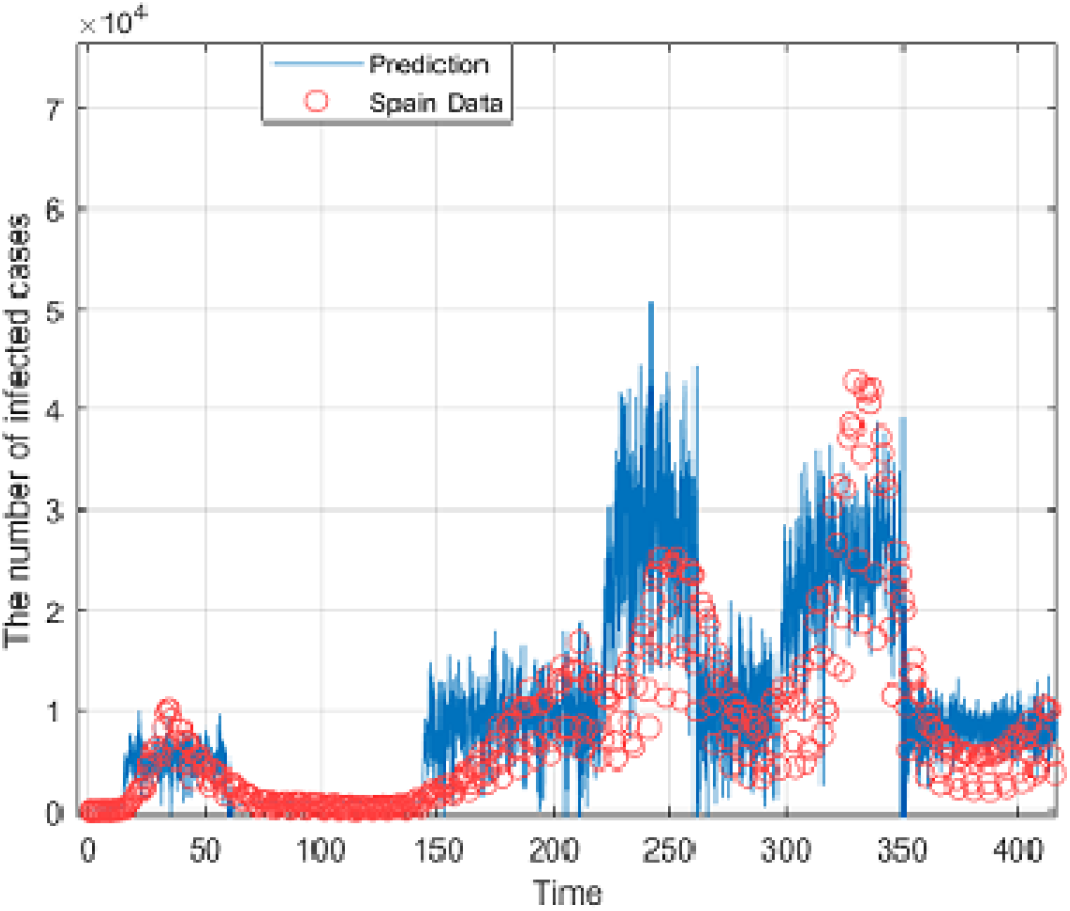
Comparison between piecewise model and real data in Spain for *α* = 1.

## 8 Conclusion

Nature can be better understood or even predicted if the limitations of existing theories, approaches, methods are questioned, revised and updated. This is also the case in epidemiological modeling, for many decades several theories have not been revisited but are still used even to analyze complex problems that cannot be really understood using existing theories. For example, how can we predict waves for a given infectious disease using existing methods? Although significant results have been suggested and some highly informative, however, when looking at the spread of Covid-19, especially data from some countries, one will quickly realize that some of them exhibit crossover behaviors, for example a passage from patterns with deterministic features to stochastic. We have attempted to open a new window of modeling such problems, by using the concept of piecewise modelling. We present some illustrative examples. The agreement of the piecewise models and experimental data let no doubt that this approach will help mankind to better predict crossover behaviors appearing in nature.

## Data Availability

Not applicable

